# Measurement of quality of stroke care with national electronic health records: a prospective cohort study during and after the COVID-19 pandemic

**DOI:** 10.64898/2025.12.03.25340732

**Authors:** James Farrell, John Nolan, Roger Lambert, Ana Torralbo, Steffen E. Petersen, Mevhibe Hovcaoglu, Chris Tomlinson, Reecha Sofat, Qi Huang, Evan Kontopantelis, Martin James, Sarah Lessels, Jacqueline A. L. MacArthur, Angela Wood, William N Whiteley, Spiros Denaxas CVD-COVID-UK/COVID-IMPACT Consortium

## Abstract

**Objectives:** To evaluate the value of linked electronic health records (EHRs) for measuring stroke care quality in England before and after the COVID-19 pandemic, focusing on metrics not routinely captured: stroke incidence, dispensing of secondary prevention medications, and a proxy of disability—time spent at home after stroke (“home-time”).

**Design:** Prospective cohort study using national linked datasets.

**Setting:** England-wide health data linkage including the Sentinel Stroke National Audit Programme (SSNAP), primary and secondary care, dispensed medications, and mortality records, accessed via NHS England’s Secure Data Environment.

**Participants:** 425,675 adults with a first stroke between 1 January 2020 and 31 December 2023; data were available for 304,210 in primary care, 279,825 in hospital admissions, 220,470 in SSNAP, and 59,465 in death records.

**Main outcome measures:** Annual stroke incidence; first-year medication dispensing rates for antiplatelets, anticoagulants, antihypertensives, and lipid-lowering agents; and home-time at 180 days post-stroke.

**Results:** Stroke ascertainment was highest when combining all sources, with 10.8% of non-fatal ischaemic strokes recorded exclusively in primary care and 19.4% of fatal strokes identified solely through death records. Standardised annual stroke incidence rose from 227.6 [95% CI 226.1, 229.0] to 244.8 [95% CI 243.4, 246.3] per 100,000 over the study period including the COVID-19 pandemic. During the COVID-19 lockdown, non-fatal stroke recordings decreased while stroke-related deaths rose, indicating that recording quality was sensitive to shifts in healthcare-seeking behavior during the pandemic. Among people with ischaemic stroke, 89.1% received an antiplatelet or anticoagulant, 44.5% an antihypertensive, and 80.5% a lipid-lowering therapy. For haemorrhagic stroke, these proportions were, for anticoagulants 13.5%, antiplatelets 13.2%, antihypertensives 46.6%, and lipid lowering 41.1%. Medication dispensing for stroke prevention declined with increasing age and comorbidity but varied little by ethnicity, region, or pandemic period. Mean home-time within 180 days of stroke was 166.6 [95% CI 166.4, 166] days, decreasing with greater age (141.4 days for 90 years or older [95% CI 140.7, 142.1]), deprivation (166.4 [95% CI 166.1, 166.6] for most deprived quintile), and stroke severity (137.4 days for NIHSS score on arrival over 22 [95% CI 135.8, 139.1]), and increasing with years from the COVID-19 pandemic 2023 169.3 days [95% CI 169.0, 169.5] vs. 2020 164.4 days [95% CI 164.4 [164.1, 164.7]).

**Conclusions:** Standardised stroke incidence increased significantly over the study period, highlighting a growing public health burden that persisted despite disruptions due to the pandemic. While secondary prevention coverage for antiplatelets and lipids was high, the suboptimal dispensing of antihypertensives, particularly in older and comorbid populations, signals a critical target for clinical optimisation. Home-time represents a sensitive, person-centered outcome that exposes disparities linked to socioeconomic deprivation and clinical severity that can be used to enhance routine stroke audits. These findings justify the expansion of linked EHR infrastructure and the modernisation of governance frameworks to enable the longitudinal evaluation of care quality beyond the COVID-19 era.

**Strengths and Limitations of this Study:**

- This study utilises a whole-population linkage of national audit (SSNAP), primary care (GDPPR), hospital (HES), and mortality (ONS) records, which maximises case ascertainment compared to routine audits.
- The use of NHS Business Services Authority (NHSBSA) medication dispensing data potentially provides a more accurate proxy for medication adherence than primary care prescribing records alone.
- The inclusion of primary care records enables capture of patients not admitted to hospital or managed in ambulatory settings, while linkage with hospital records allows longitudinal measurement of ‘home-time’ following stroke.
- Potential misclassification of stroke subtypes and first-ever versus recurrent stroke events remains a risk due to the varying PPV of ICD-10 and SNOMED-CT concepts in electronic health records from different healthcare settings.
- The analysis is unable to account for medications prescribed in the private sector or clinical factors not captured in routine electronic health records, such as specific reasons for contraindications, prior tolerance, or eligibility to secondary prevention.

## Introduction

Near-real-time linkage of routine health and medication dispensing records can enhance national stroke audits by improving case ascertainment and reducing the time and cost of data collection. Furthermore, integrating this data allows for the identification of care inequalities and the development of new quality measures, such as six-month disability indicators and secondary prevention coverage.Here we test the benefits of linking the Sentinel Stroke National Audit Programme (SSNAP) in England to wider health-systems data during the COVID-19 pandemic, when quality of care was expected to vary substantially [1].

UK national audits of stroke care like SSNAP measure quality of care for people admitted to hospitals with stroke. In addition, audits provide high fidelity data on particular diseases that are otherwise not available in UK hospitalisation or death records, hence are particularly useful for public health and epidemiological stroke research. SSNAP makes considerable effort to ensure complete and accurate ascertainment of people admitted to hospitals. This is resource intensive, but despite these efforts, audits may not ascertain everybody affected - for example, the Acute Myocardial Ischaemia National Audit Project did not record myocardial infarction cases primarily ascertained in primary care (for example from emergency department discharges) and at death [2]. It is possible that stroke audits also miss some people affected by stroke - for example those with milder stroke who are not admitted to hospital, people who are looked after outside of a stroke service, or those who die very quickly in emergency departments - and may be particularly affected by health emergencies such as the COVID-19 pandemic.

Here, we used linked national electronic health records—including SSNAP, primary care, hospital admissions, dispensed medications, and mortality data—to evaluate how stroke ascertainment varied across sources, to assess the impact of the COVID-19 pandemic on stroke recording and care, and to test whether routinely collected data can support quality measurement across three domains: completeness of case capture across datasets to inform future surveillance approaches; implementation of secondary prevention with medication dispensing, including antiplatelets, antihypertensives, and statins; and use of “home-time” in the first six months post-stroke as a proxy for functional outcome.

## Methods

### Study design and data sources

#### Study population

We implemented a prospective cohort study and identified people over 18 years with a diagnosis of stroke between 1st January 2020 and 31st December 2023 and who were registered with a general practitioner (GP), living in England, had at least one record in General Practice Extraction Service Data for Pandemic Planning and Research (GDPPR) and had a valid pseudo-identifier [3]. Residents of England were identified with the lower layer super output area (LSOA) of residence.

We linked each person to Hospital Episode Statistics (HES), GDPPR, medication dispensing in primary care (NHS Business Services Authority (NHSBSA))[4], and Office for National Statistics death registrations (ONS Deaths) [5]. Data were accessed via the BHF Data Science Centre’s CVD-COVID-UK/COVID-IMPACT Consortium in NHS England’s SDE service for England [3]. Individual data sources were linked by NHS Digital using the Master Person Service in combination with the Personal Demographics Service and stringent data quality controls were applied upstream by NHS Digital to ensure data linkages were accurate.

#### Identifying strokes

The first stroke event (index stroke) was defined as the earliest record across multiple sources: hospital admissions in SSNAP or HES–Admitted Patient Care (APC), primary care records in GDPPR, or the date of death in ONS Deaths. Stroke records from different sources were considered the same event if their dates fell within 30 days of the index stroke date, to allow for minor discrepancies in recording. Individuals with any stroke records in these sources prior to the study start date were excluded from the analyses.

We classified index strokes into three mutually exclusive phenotypes: ischaemic stroke, haemorrhagic stroke, and unknown stroke type. Stroke type was determined using different data sources. In SSNAP, stroke type was recorded in the field “S2 stroke type.” In HES-APC and ONS Deaths, stroke was identified using ICD-10 codes, I63 for ischaemic stroke (cerebral infarction), I61 for haemorrhagic stroke (intracerebral haemorrhage), and I64 for unknown stroke type (not specified as haemorrhage or infarction). In HES-APC, we used the ICD-10 from the primary diagnosis of any episode within a spell, and in ONS Deaths, from the cause of death. In GDPPR, stroke was identified and classified with SNOMED-CT concepts as outlined in the supplement.To consolidate stroke type across sources, we prioritised records in the following order: SSNAP, HES-APC, GDPPR, and ONS Deaths. If a source with the highest priority contained both ischaemic and haemorrhagic stroke records, the stroke type was classified as unknown. Index strokes were further classified as fatal (death from any cause within 30 days) or non-fatal.

#### Covariate definitions

Age, sex, and ethnicity were obtained from the most recent non-missing value across primary care (GDPPR) and secondary care (HES-APC). When values conflicted between sources, priority was given to primary care. Ethnicity was classified in the ONS categories: White, Asian or Asian British, Black or Black British, Mixed, or Other [6].

Socioeconomic deprivation was derived by linking the 2011 LSOAs to the 2019 Index of Multiple Deprivation (IMD) and stratified into quintiles from most to least deprived.

GDPPR and HES-APC (primary and non-primary diagnoses) were used to define prevalent conditions between birth and a day prior to the earliest stroke: atrial fibrillation (AF), cancer, dementia, type 2 diabetes (DM), hypertension, obesity, chronic obstructive pulmonary disease (COPD), stable angina, depression, deep vein thrombosis (DVT) and chronic kidney disease (CKD). Hypercholesterolaemia was defined with GDPPR alone. COVID-19 infection was defined with HES-APC, GDPPR, and COVID-19 testing data (Second Generation Surveillance System (SGSS) and the UK Non-hospital COVID-19 testing dataset (also known as Pillar 2) [7].

We defined smoking status, body mass index (BMI), systolic blood pressure (SBP), estimated glomerular filtration rate (eGFR), glycated haemoglobin (HbA1c), total cholesterol and high density lipoprotein cholesterol (HDL) by extracting SNOMED concepts using previously validated phenotyping [7] algorithms from GDPPR between two years and one day prior to the earliest recorded stroke and within predefined value ranges (supplied in Appendix).

#### Outcome definitions

Home-time (or time out of hospital) was defined as days alive and not in hospital in the 180 days from first stroke record date, where hospital time was defined as the sum of the duration of all admission to discharge spells (or end of follow up) within the first six months. We defined mortality as death of any cause within 30 days of the first recorded stroke. We extracted relevant dispensed medications (e.g. statins, antihypertensives, anticoagulants, and antiplatelets) from NHSBSA using previously validated phenotyping algorithms [8]. Previous medications were defined as prescriptions in the year prior to the earliest stroke event. Incident prescriptions were defined as a prescription from one month after discharge date (for hospitalised stroke) or earliest stroke date (if not hospitalised) from one month to one year of the index event.

Phenotyping algorithms, codelists, and related metadata are provided in the Supplementary Materials (Table S6).

### Statistical analyses

We used descriptive statistics to summarise patient populations and characteristics in stroke that were common or unique to each data source. For the earliest stroke, we plot and report the differences between event dates recorded in HES-APC, GDPPR and SSNAP.

We used UpSet plots to describe the agreement between initial stroke records in GDPPR, HES-APC, SSNAP and ONS Deaths. Crude and adjusted mean home-time was calculated for the overall cohort and stratified by stroke type, demographics, comorbidities, data source, and for patients in SSNAP, NIHSS on arrival, and modified Rankin score at discharge. Both unadjusted and adjusted estimates were reported with a multivariable linear model.

The one-year cumulative incidence of post-stroke dispensed medications for anticoagulants, antiplatelets, antihypertensives, and lipid-lowering drugs was calculated, with death as a competing event. Monthly stroke incidence rates between 2020 and 2023 where age and sex standardised to the European Standard Population (ESP) using the direct method.

Cumulative incidence was stratified by demographic factors, Charlson comorbidity index, data source, atrial fibrillation, and pre-stroke medication use.

A multivariable cause-specific Cox proportional hazards model was used to estimate hazard ratios for covariates and to calculate the covariate-adjusted cumulative incidence of first post-stroke dispensed medication. Formal evaluation of the proportional hazards assumption was not performed, as the estimated hazard ratios were intended to summarise time-averaged effects over the follow-up period rather than to explore time-varying associations. We conducted complete data analysis.

### Data access and tools

The data used in this study are available in NHS England’s Secure Data Environment (SDE) service for England, but as restrictions apply they are not publicly available (https://digital.nhs.uk/services/secure-data-environment-service). The CVD-COVID-UK/COVID-IMPACT programme, led by the BHF Data Science Centre (https://bhfdatasciencecentre.org/), received approval to access data in NHS England’s SDE service for England from the Independent Group Advising on the Release of Data (IGARD) (https://digital.nhs.uk/about-nhs-digital/corporate-information-and-documents/independent-group-advising-on-the-release-of-data) via an application made in the Data Access Request Service (DARS) Online system (ref. DARS-NIC-381078-Y9C5K) (https://digital.nhs.uk/services/data-access-request-service-dars/dars-products-and-services). The CVD-COVID-UK/COVID-IMPACT Approvals & Oversight Board (https://bhfdatasciencecentre.org/areas/cvd-covid-uk-covid-impact/) subsequently granted approval to this project to access the data within NHS England’s SDE service for England. The de-identified data used in this study were made available to accredited researchers only. Those wishing to gain access to the data should contact in the first instance. The plan, phenotype definitions and code for this analysis are published on GitHub (https://github.com/BHFDSC/CCU005_08).

### Ethical approval

The North East-Newcastle and North Tyneside 2 research ethics committee provided ethical approval for the CVD-COVID-UK/COVID-IMPACT research programme (REC no.

20/NE/0161) to access, within secure trusted research environments, unconsented, whole-population, de-identified data from electronic health records collected as part of patients’ routine healthcare. The study was reported referencing the RECORD statement (Table S7).

### Patient and public involvement

A panel of six patient and public representatives directly affected by stroke or cardiovascular disease provided perspective and input on this research proposal, plain English summary, and research outputs, with R.L. contributing as co-author of this manuscript.

## Results

Between 1st January 2020 and 31st December 2023, we identified 425,675 people with a first stroke of whom 304,210 (71.5%) had a record in primary care, 279,825 (65.7%) in hospital admissions, 220,470 (51.8%) in SSNAP, and 59,465 (14.0%) in ONS death records, with median ages of 74.0, 76.0, 76.0, and 83.0 years respectively. A small majority of strokes in GDPPR, HES, SSNAP were in men (53.5%, 52.5%, 52.9%, respectively) and in death records, a small majority were in women (55.4%). Distribution of ethnicity, region and deprivation were similar between sources. Where the stroke was identified in death records, patients tended to be less obese, with worse eGFR, lower cholesterol and systolic blood pressure, with a greater proportion affected by comorbidities (particularly AF, chronic kidney disease, cancer and dementia) than in other sources. (Table 1)

**Table 1.** Patient characteristics, demographics and stroke-related data for individuals with a first stroke between 2020 and 2023, stratified by the presence of a stroke record in the General Practice Extraction Service Data for Pandemic Preparedness (GDPPR), Hospital Episode Statistics Admitted Patient Care (HES-APC), Sentinel Stroke National Audit Programme (SSNAP), or Office for National Statistics (ONS) Deaths. The denominator for the top row is the total number of individuals across all sources (n= 425,675); for subsequent rows, denominators are the number of individuals recorded in each data source (304,210 in GDPPR, 279,825 in HES-APC, 220,470 in SSNAP, and 59,465 in ONS).

|  |  | GDPPR | HES-APC | SSNAP | ONS Deaths | All sources |
| --- | --- | --- | --- | --- | --- | --- |
| **Individuals, n (% all sources)** |  | 304210 (71.5) | 279825 (65.7) | 220470 (51.8) | 59465 (14.0) | 425675 (100.0) |
| **Stroke type** | Ischaemic | 216295 (71.1) | 237600 (84.9) | 193680 (87.8) | 28990 (48.8) | 296730 (69.7) |
|  | Haemorrhagic | 30070 (9.9) | 38760 (13.9) | 26700 (12.1) | 15270 (25.7) | 55490 (13.0) |
|  | Unknown | 57845 (19.0) | 3460 (1.2) | 90 (0.0) | 15205 (25.6) | 73455 (17.3) |
| **Age (years)** | Median (IQR) | 74.0 (62.3 to 82.6) | 76.0 (64.6 to 84.4) | 76.0 (64.8 to 84.3) | 83.0 (74.2 to 89.5) | 75.5 (63.8 to 84.1) |
| **Age group (years)** | 18-59 | 63760 (21.0) | 49335 (17.6) | 38165 (17.3) | 4660 (7.8) | 80300 (18.9) |
|  | 60-69 | 58080 (19.1) | 48110 (17.2) | 38255 (17.4) | 5825 (9.8) | 74670 (17.5) |
|  | 70-79 | 84295 (27.7) | 75410 (26.9) | 60080 (27.3) | 13425 (22.6) | 114515 (26.9) |
|  | 80-89 | 74915 (24.6) | 78835 (28.2) | 62360 (28.3) | 21740 (36.6) | 114665 (26.9) |
|  | 90+ | 23160 (7.6) | 28130 (10.1) | 21615 (9.8) | 13815 (23.2) | 41520 (9.8) |
| **Sex** | Male | 162830 (53.5) | 146890 (52.5) | 116720 (52.9) | 26500 (44.6) | 222245 (52.2) |
| **Ethnicity** | White | 272065 (89.4) | 251220 (89.8) | 198745 (90.1) | 54595 (91.8) | 380625 (89.4) |
|  | Asian | 16885 (5.6) | 14895 (5.3) | 11495 (5.2) | 2390 (4.0) | 23380 (5.5) |
|  | Black | 8715 (2.9) | 7990 (2.9) | 6000 (2.7) | 1180 (2.0) | 12375 (2.9) |
|  | Mixed | 2425 (0.8) | 2080 (0.7) | 1540 (0.7) | 335 (0.6) | 3325 (0.8) |
|  | Other | 3150 (1.0) | 2610 (0.9) | 1920 (0.9) | 435 (0.7) | 4310 (1.0) |
|  | Unknown | 970 (0.3) | 1030 (0.4) | 775 (0.4) | 535 (0.9) | 1660 (0.4) |
| **Region** | East Midlands | 28675 (9.4) | 25515 (9.1) | 20785 (9.4) | 5385 (9.1) | 38855 (9.1) |
|  | East of England | 33950 (11.2) | 29935 (10.7) | 24390 (11.1) | 6265 (10.5) | 45900 (10.8) |
|  | London | 32830 (10.8) | 30555 (10.9) | 22245 (10.1) | 5925 (10.0) | 48445 (11.4) |
|  | North East | 18530 (6.1) | 16725 (6.0) | 13930 (6.3) | 3030 (5.1) | 24200 (5.7) |
|  | North West | 39855 (13.1) | 40050 (14.3) | 32620 (14.8) | 8310 (14.0) | 59650 (14.0) |
|  | South East | 49600 (16.3) | 44600 (15.9) | 33765 (15.3) | 10145 (17.1) | 69700 (16.4) |
|  | South West | 37025 (12.2) | 31910 (11.4) | 26845 (12.2) | 7325 (12.3) | 49650 (11.7) |
|  | West Midlands | 29330 (9.6) | 29805 (10.7) | 22610 (10.3) | 6640 (11.2) | 43210 (10.2) |
|  | Yorkshire and The Humber | 34410 (11.3) | 30730 (11.0) | 23280 (10.6) | 6445 (10.8) | 46070 (10.8) |
| **IMD 2019 quintiles** | 1 (most deprived) | 58875 (19.4) | 56045 (20.0) | 44100 (20.0) | 11390 (19.2) | 84165 (19.8) |
|  | 2 | 59640 (19.6) | 55405 (19.8) | 43405 (19.7) | 11580 (19.5) | 84125 (19.8) |
|  | 3 | 62700 (20.6) | 57305 (20.5) | 45050 (20.4) | 12480 (21.0) | 87370 (20.5) |
|  | 4 | 62920 (20.7) | 57145 (20.4) | 45130 (20.5) | 12345 (20.8) | 87210 (20.5) |
|  | 5 (least deprived) | 60070 (19.7) | 53925 (19.3) | 42785 (19.4) | 11670 (19.6) | 82805 (19.5) |
| **Smoking status** | Current smoker | 40105 (13.2) | 34845 (12.5) | 27510 (12.5) | 5445 (9.2) | 53210 (12.5) |
|  | Ex-smoker | 76770 (25.2) | 70305 (25.1) | 55705 (25.3) | 16030 (27.0) | 107990 (25.4) |
|  | Never smoked | 92825 (30.5) | 83375 (29.8) | 65470 (29.7) | 18695 (31.4) | 129430 (30.4) |
|  | Unknown | 94515 (31.1) | 91305 (32.6) | 71790 (32.6) | 19290 (32.4) | 135040 (31.7) |
| **Body mass index (BMI)** | Median (IQR) | 27.3 (23.9 to 31.2) | 27.1 (23.7 to 31.2) | 27.2 (23.8 to 31.2) | 25.1 (21.6 to 29.2) | 27.0 (23.6 to 31.0) |
| **eGFR** | Median (IQR) | 70.0 (56.0 to 83.0) | 67.0 (53.0 to 82.0) | 67.0 (53.0 to 81.0) | 62.0 (47.0 to 78.0) | 68.0 (54.0 to 82.0) |
| **Glycated haemoglobin** | Median (IQR) | 41.0 (37.0 to 48.0) | 41.0 (38.0 to 49.0) | 41.0 (38.0 to 48.0) | 41.0 (37.0 to 47.0) | 41.0 (37.0 to 48.0) |
| **Systolic blood pressure** | Median (IQR) | 136.0 (126.0 to 148.0) | 137.0 (126.0 to 148.0) | 137.0 (126.0 to 148.0) | 133.0 (120.0 to 145.0) | 136.0 (125.0 to 147.0) |
| **HDL cholesterol** | Median (IQR) | 1.3 (1.1 to 1.6) | 1.3 (1.1 to 1.6) | 1.3 (1.1 to 1.6) | 1.3 (1.1 to 1.7) | 1.3 (1.1 to 1.6) |
| **Total cholesterol** | Median (IQR) | 3.4 (2.7 to 4.2) | 3.3 (2.7 to 4.2) | 3.3 (2.7 to 4.2) | 3.1 (2.5 to 3.9) | 3.3 (2.7 to 4.2) |
| **Angina** | Yes | 48370 (15.9) | 48665 (17.4) | 38340 (17.4) | 12970 (21.8) | 72480 (17.0) |
| **Arrhythmia** | Yes | 4465 (1.5) | 5000 (1.8) | 3395 (1.5) | 2545 (4.3) | 8150 (1.9) |
| **Atrial fibrillation or myocardial infarction** | Yes | 90040 (29.6) | 99525 (35.6) | 77400 (35.1) | 29785 (50.1) | 141030 (33.1) |
| **Diabetes** | Yes | 88280 (29.0) | 86050 (30.8) | 66960 (30.4) | 19050 (32.0) | 127320 (29.9) |
| **Hypercholesterolaemia** | Yes | 25965 (8.5) | 24800 (8.9) | 19550 (8.9) | 5580 (9.4) | 37515 (8.8) |
| **Hypertension** | Yes | 207235 (68.1) | 203950 (72.9) | 159755 (72.5) | 46645 (78.4) | 299035 (70.2) |
| **Obesity** | Yes | 61395 (20.2) | 58675 (21.0) | 45415 (20.6) | 11150 (18.7) | 86280 (20.3) |
| **Cancer** | Yes | 72570 (23.9) | 69470 (24.8) | 53720 (24.4) | 18980 (31.9) | 106920 (25.1) |
| **Chronic kidney disease** | Yes | 87555 (28.8) | 93005 (33.2) | 70600 (32.0) | 30465 (51.2) | 139235 (32.7) |
| **Liver disease** | Yes | 4405 (1.4) | 4230 (1.5) | 3085 (1.4) | 1360 (2.3) | 6910 (1.6) |
| **Chronic obstructive pulmonary disease** | Yes | 39305 (12.9) | 39490 (14.1) | 30650 (13.9) | 11135 (18.7) | 59630 (14.0) |
| **Dementia** | Yes | 22675 (7.5) | 25310 (9.0) | 18875 (8.6) | 13765 (23.1) | 41330 (9.7) |
| **Depression** | Yes | 83235 (27.4) | 73255 (26.2) | 57025 (25.9) | 15575 (26.2) | 115745 (27.2) |
| **Deep vein thrombosis** | Yes | 7640 (2.5) | 7965 (2.8) | 5835 (2.6) | 2720 (4.6) | 12525 (2.9) |
| **Charlson comorbidity index** | 0 | 60850 (20.0) | 23700 (8.5) | 24310 (11.0) | 4610 (7.8) | 70635 (16.6) |
|  | 1-2 | 115525 (38.0) | 106585 (38.1) | 83595 (37.9) | 15690 (26.4) | 154310 (36.3) |
|  | 3-4 | 73680 (24.2) | 81880 (29.3) | 62610 (28.4) | 16720 (28.1) | 108580 (25.5) |
|  | 5+ | 54155 (17.8) | 67655 (24.2) | 49955 (22.7) | 22445 (37.7) | 92150 (21.6) |
| **COVID-19 within 14 days prior to** | Yes | 8230 (2.7) | 10110 (3.6) | 6475 (2.9) | 3825 (6.4) | 14965 (3.5) |
| **stroke** |  |  |  |  |  |  |
| **History of COVID-19** | Yes | 45765 (15.0) | 42980 (15.4) | 31445 (14.3) | 12625 (21.2) | 69150 (16.2) |
| **Pre-stroke anticoagulants** | Yes | 45645 (15.0) | 48925 (17.5) | 38260 (17.4) | 16140 (27.1) | 72605 (17.1) |
| **Pre-stroke antihypertensives** | Yes | 121915 (40.1) | 115805 (41.4) | 91845 (41.7) | 24240 (40.8) | 172175 (40.4) |
| **Pre-stroke antiplatelets** | Yes | 103560 (34.0) | 82200 (29.4) | 66300 (30.1) | 16465 (27.7) | 138315 (32.5) |
| **Pre-stroke lipid lowering drugs** | Yes | 155180 (51.0) | 136375 (48.7) | 109190 (49.5) | 26785 (45.0) | 212980 (50.0) |
| **Length of stay (days)** | Median (IQR) | 4.0 (2.0 to 12.0) | 5.0 (2.0 to 16.0) | 5.0 (2.0 to 14.0) | 6.0 (2.0 to 12.0) | 5.0 (2.0 to 15.0) |
| **NIHSS score on arrival** | 0-4 | 89305 (29.4) | 106580 (38.1) | 116295 (52.7) | 3575 (6.0) | 116295 (27.3) |
|  | 5-10 | 38520 (12.7) | 49950 (17.9) | 53335 (24.2) | 3580 (6.0) | 53335 (12.5) |
|  | 11-15 | 11510 (3.8) | 17230 (6.2) | 18370 (8.3) | 3155 (5.3) | 18370 (4.3) |
|  | 16-21 | 9670 (3.2) | 16680 (6.0) | 17655 (8.0) | 5510 (9.3) | 17655 (4.1) |
|  | 22+ | 6120 (2.0) | 13615 (4.9) | 14680 (6.7) | 7650 (12.9) | 14680 (3.4) |
|  | (missing) | 149085 (49.0) | 75770 (27.1) | 140 (0.1) | 35990 (60.5) | 205345 (48.2) |
| **Rankin score at discharge** | 0 | 15660 (5.1) | 17800 (6.4) | 19755 (9.0) | 135 (0.2) | 19755 (4.6) |
|  | 1 | 25930 (8.5) | 29555 (10.6) | 32055 (14.5) | 135 (0.2) | 32055 (7.5) |
|  | 2 | 20820 (6.8) | 23510 (8.4) | 25335 (11.5) | 175 (0.3) | 25335 (6.0) |
|  | 3 | 16440 (5.4) | 19375 (6.9) | 20690 (9.4) | 325 (0.5) | 20690 (4.9) |
|  | 4 | 12865 (4.2) | 16635 (5.9) | 17715 (8.0) | 740 (1.2) | 17715 (4.2) |
|  | 5 | 5720 (1.9) | 8350 (3.0) | 8895 (4.0) | 2055 (3.5) | 8895 (2.1) |
|  | 6 | 3960 (1.3) | 18015 (6.4) | 20160 (9.1) | 17145 (28.8) | 20160 (4.7) |
|  | (missing) | 202815 (66.7) | 146580 (52.4) | 75870 (34.4) | 38765 (65.2) | 281070 (66.0) |

The estimated incidence of stroke during the period 1st January 2020 and 31st December 2023 was highest for a comprehensive ascertainment of stroke from all sources, compared with individual sources alone. The incidence rate per 100,000 person years, age- and sex-standardised to the European Standard Population, for strokes identified from all four sources was 227.6 [226.1, 229.0] in 2020 and rose to 244.8 [243.4, 246.3] in 2023 (Table S1).

The percentage of people with ischaemic, haemorrhagic, and unknown stroke types was, in primary care 71.1%, 9.9%, 19.0%, in hospital statistics 84.9%, 13.9%, 1.2%, in SSNAP 87.8%, 12.1%, <0.1%, and in death records 48.8%, 25.7%, 25.6% respectively. Using the harmonised stroke type, these proportions were 69.7%, 13.0% and 17.3%. (Table 1)

Non-fatal ischaemic strokes in SSNAP were almost always recorded in HES-APC as ischaemic or unknown and were rarely recorded without at least one other corroborating source (3.6% of non-fatal strokes). Haemorrhagic strokes in SSNAP were very rarely recorded without another corroborating source (0.6% of non-fatal strokes) and were almost always recorded as haemorrhagic in primary or hospital records. There were a significant number of patients with a diagnosis of ischaemic stroke who were only recorded in primary care (10.8% of non-fatal strokes) or hospital records (4.6% of non-fatal strokes), and the largest number of haemorrhagic strokes were only recorded in primary care (2.0% of non-fatal strokes). (Figure 1). The largest number of fatal strokes (19.4%) had no subtype information and were only recorded in death records. Death records were the only source of subtype information for 4,620 haemorrhagic strokes (6.8% of fatal strokes) and 3,515 ischaemic strokes (5.2% of fatal strokes).(Table S2a-b, Figure 1)

**Figure 1.**
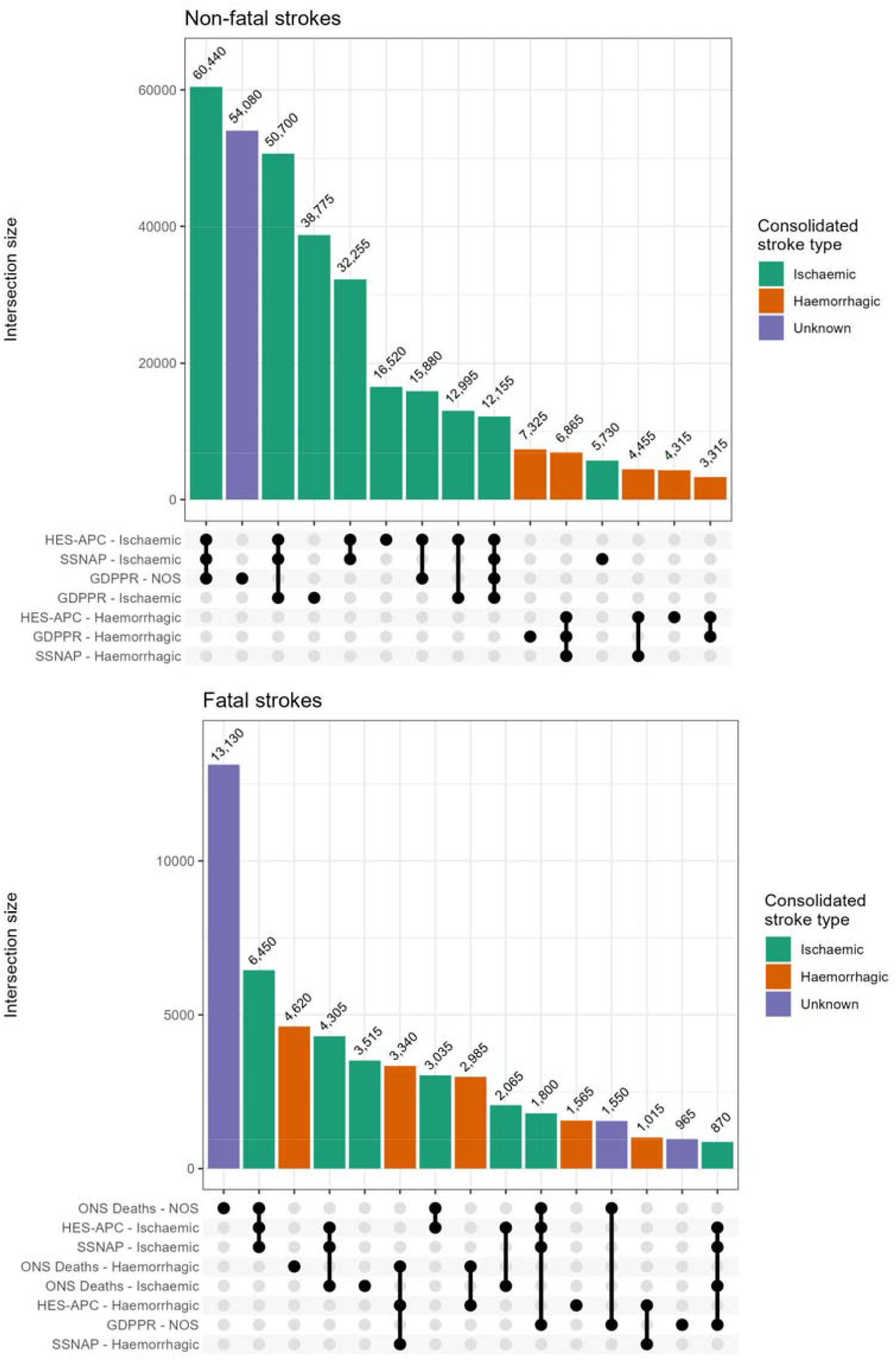
UpSet plot of the 15 most frequent intersections of stroke events across GDPPR, HES-APC, SSNAP, and ONS Deaths by stroke type. Counts represent individuals with non-fatal (n=358,025) and fatal strokes (n=67,650). NOS: not otherwise specified.

The median difference in stroke onset date between sources, compared from the earliest recorded date within each source, was modest. Between primary care and hospital episodes it was 0 days (IQR 0-4), primary care and SSNAP 0 days (IQR 0-4), SSNAP and hospital episodes 0 days (IQR 0-0) (Figure S1).

The highest incidence of stroke in an individual source for calendar year 2023 was recorded in primary care at 172.6 [171.3, 173.8] per 100,000, followed by hospital episodes (160.5 [159.4, 161.7]), SSNAP (128.2 [127.2, 129.3]), then death records (31.1 [30.6, 31.7]). During early 2020 (the lockdown period of the COVID-19 pandemic), non-fatal strokes recorded during healthcare encounters fell, although deaths due to stroke rose. During the pandemic, incidence of stroke classified as ischaemic fell by more than strokes classified as unknown or haemorrhagic (Figure 2).

**Figure 2.**
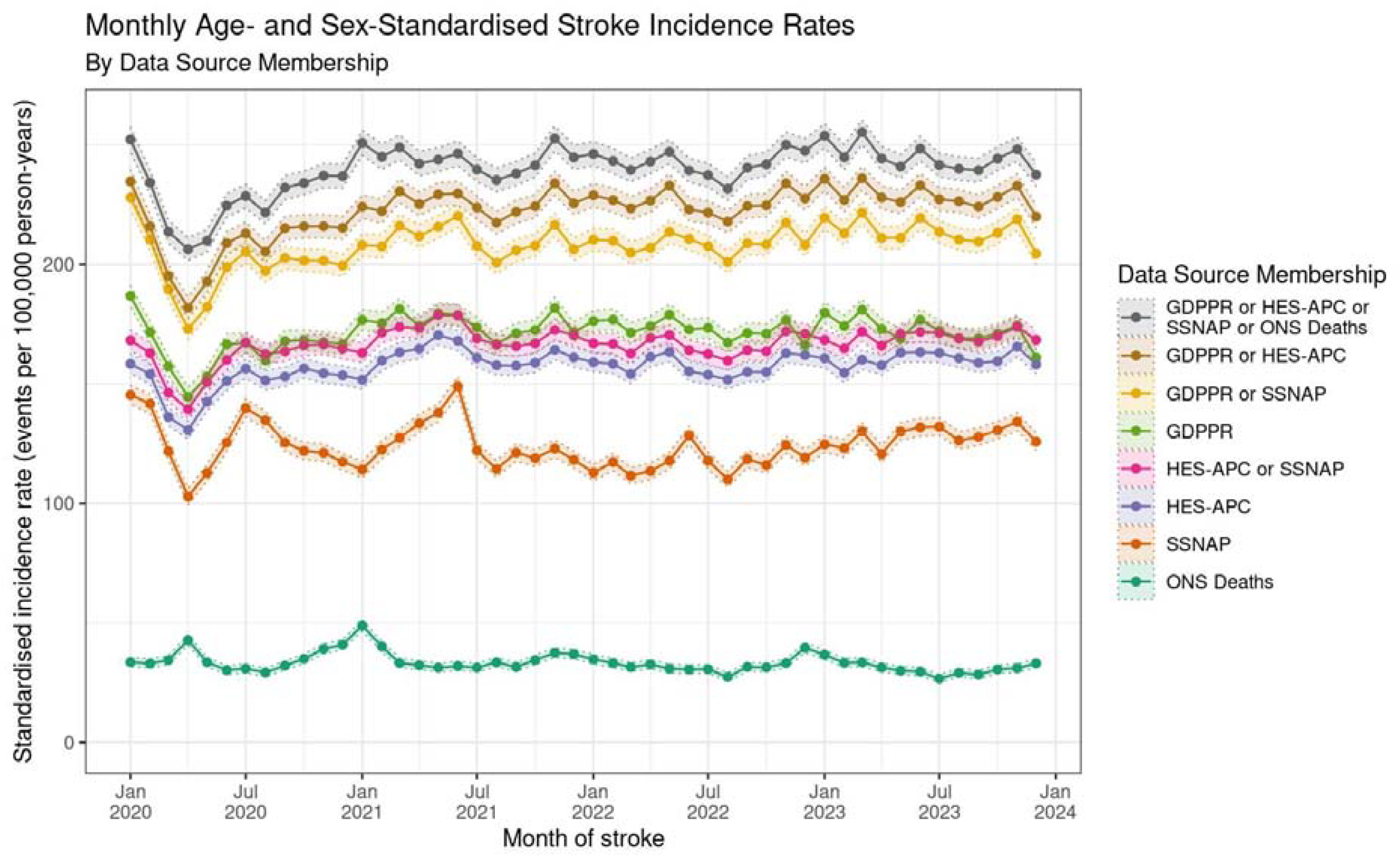
Monthly age- and sex-standardised incidence rates of stroke (standardised to the European Standard Population) between 2020 to 2023 for individuals aged 18 years and older by data source. COVID-19 lockdowns were March 2020 to July 2020, and November 2020 to December 2020.

For ischaemic stroke, within the first year of stroke, the proportion of people dispensed an: antiplatelet or anticoagulant was 89.1%, anticoagulant medication was 27.0%, antiplatelet medication was 68.9%, antihypertensive medication 44.5%, and lipid lowering medications 80.5% (Figure 3). For haemorrhagic stroke, the proportion of people dispensed a prescription of anticoagulant medication within the first year was 13.5%, antiplatelet medication was 13.2%, antihypertensive medication 46.6%, and lipid lowering medications 41.1% (Figure 3).

**Figure 3.**
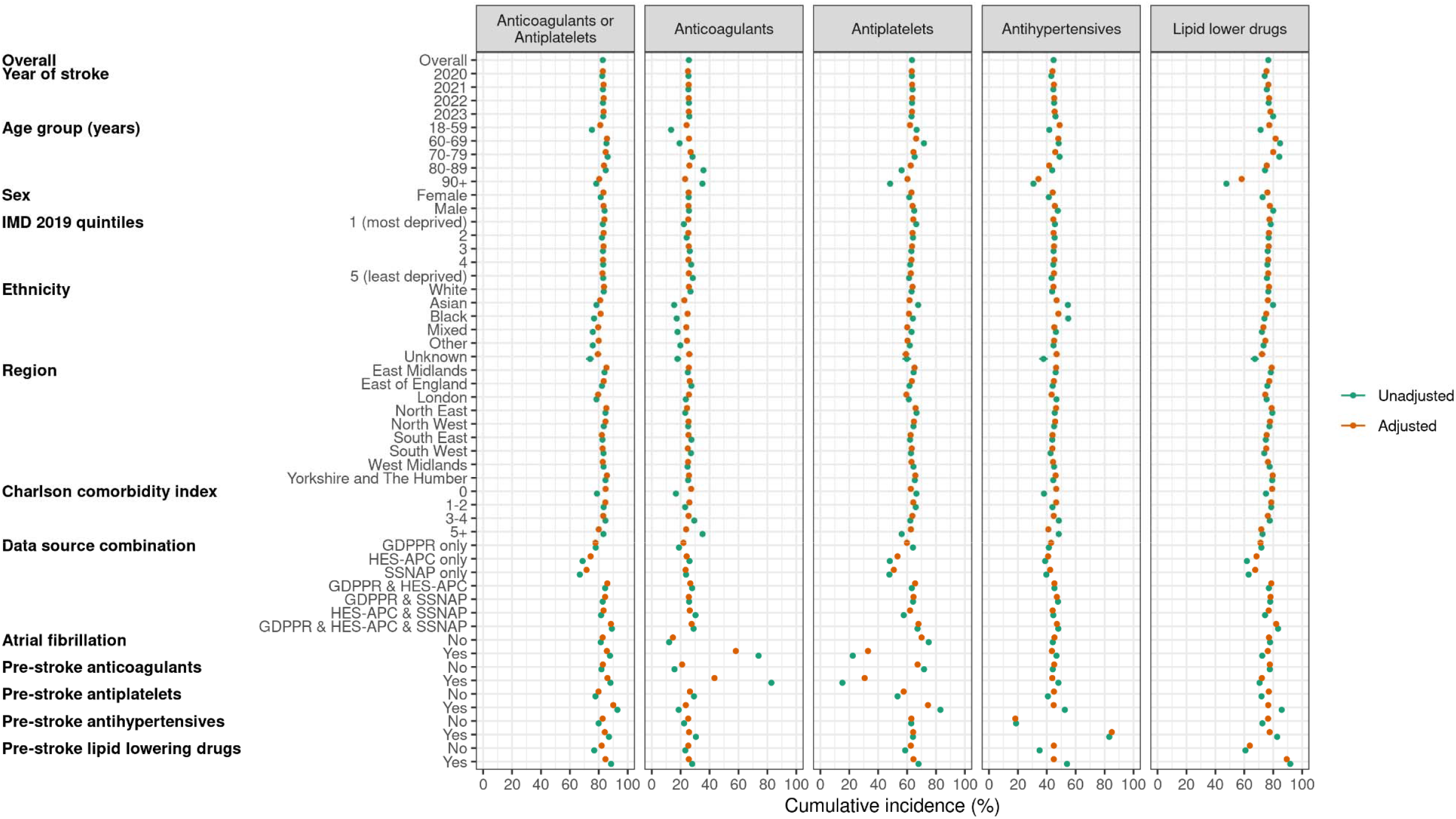
Unadjusted and covariate-adjusted one-year cumulative incidence of anticoagulants, antiplatelets, antihypertensives, and lipid-lowering drugs dispensed to individuals with non-fatal strokes identified through GDPPR, HES-APC, or SSNAP (n=348,300).

After adjusting for other stroke characteristics, older people were less likely to be dispensed lipid lowering or antihypertensive medication but not anticoagulants or antiplatelets, with no large differences by ethnicity or consistent differences for different drug classes by region. People with the greatest number of comorbidities were less likely to be dispensed any of the medicine classes than people with fewer comorbidities, although the absolute differences were small. 77.1% of people with prior atrial fibrillation and ischaemic stroke were dispensed an anticoagulant in the following year. For years 2020 to 2023, the 1-year cumulative incidence of both anticoagulants and antiplatelets dispensed was similar at 25% and 63% respectively, whilst antihypertensives increased from 43.1% to 46.0% and lipid-lowering drugs from 74.1% to 79.9% (Supplementary tables 4a-c). There was no important difference in 2020 compared to 2021 to 2023.

The mean home-time in the first 180 days post-stroke was 166.6 days, with good evidence of less home-time with increasing age, deprivation, comorbidity and NIHSS score on arrival and important differences by region in crude and adjusted analysis (Table S4.a). Home-time within 180 days post-stroke increased with time since the COVID-19 pandemic from 164.4 days (2020) to 165.5 days (2021), 166.8 days (2022), and 169.3 days (2023) (Figure 4, Table S5.a)

**Figure 4.**
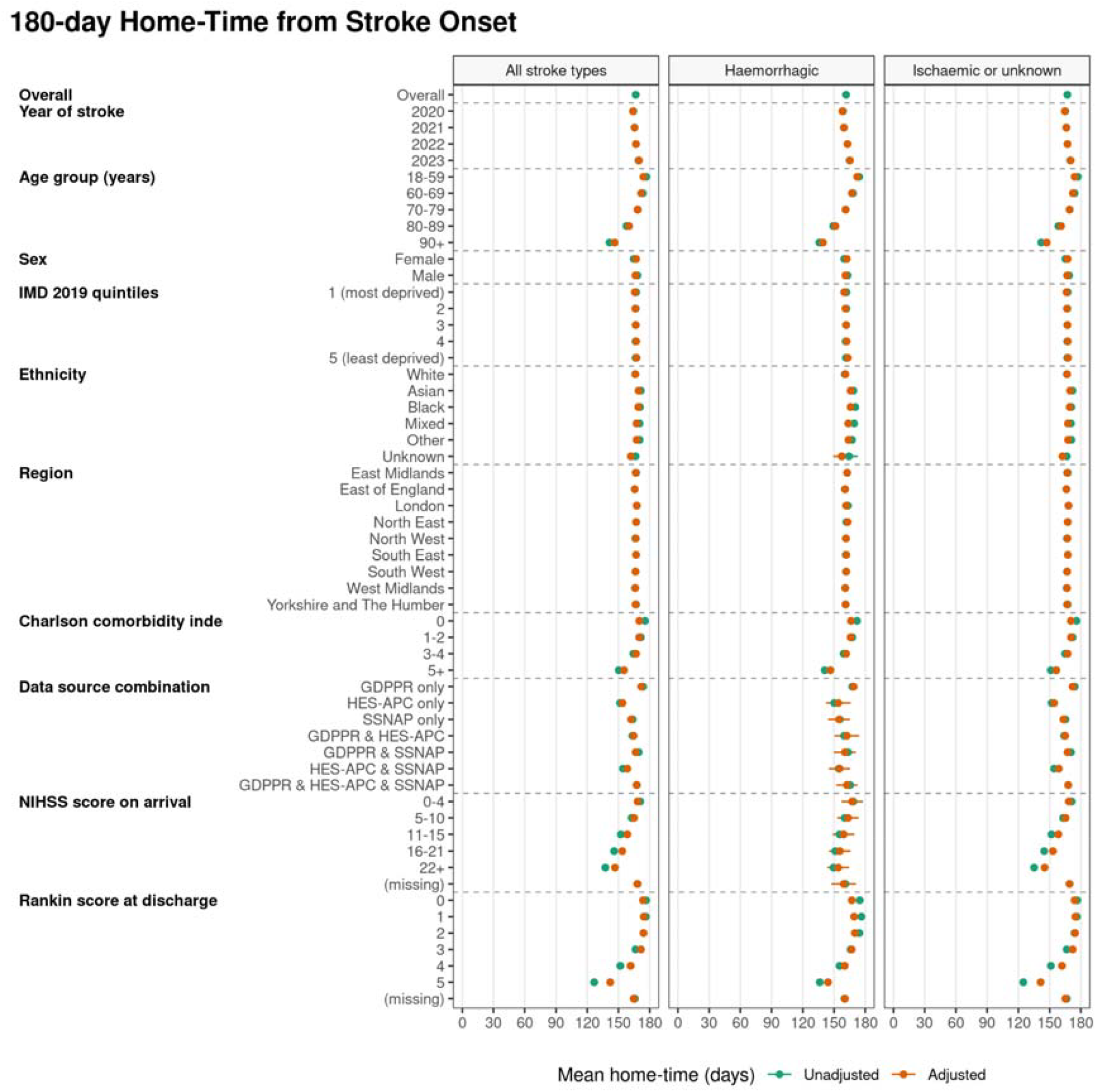
Unadjusted and covariate-adjusted mean 180-day home-time from stroke onset in non-fatal strokes identified through GDPPR, HES-APC, or SSNAP, stratified by all stroke types (n=353,325), hemorrhagic strokes (n=35,165), and ischemic or unknown stroke types (n=318,160).

After adjusting for other characteristics (age, comorbidities and NIHSS), the numbers of days of home-time in 180 days after stroke onset was fewer: compared with patients who were 15-59 years old, for patients 60-69 (by 2.1 days); 70-79, (by 5.6 days); 80-89, (by 13.8 days); and 90+, (by 27.6 days). Compared with women, men had 0.6 fewer days at home; compared with people in the least deprived fifth, those in the most deprived fifth had 1.6 days fewer at home; compared with those with no comorbidities, those with a Charlson score of 5 or more had 14.7 days less at home; and compared with those with an NIHSS score of 0-4 (mild stroke) the number of days fewer at home was, for people with an NIHSS score of 5-10, 3.3 days; 11-15, 9.9 days; 16-21, 14.7 days; and more than 22 was 21.7 days. (Table S5.b). Compared with people with a white ethnicity, the numbers of days more at home was, for those: with an Asian ethnicity, 3.0 days; with a Black ethnicity 2.8 days; those with a mixed ethnicity 1.1 days, those with other ethnicity 1.2 days, and for those with an unknown ethnicity, 4.4 fewer days at home. There were modest differences in home-time between regions, with, compared with the East Midlands, people in the East of England spending 0.9 fewer days at home, and people in London spending 0.9 days more at home. Compared with 2020, the number of days at home was 5.8 days more in 2023. (Figure 4, Table S5.b).

## Discussion

While each individual data source captured stroke events that were uniquely recorded in that source, the overall concordance of stroke events between sources was very high (median 0 days between records) indicating both a good level of data linkage across sources and quality within sources. We additionally demonstrate, with a comprehensive ascertainment of stroke in all available sources in England, that during the study period (2020-2023) most ischaemic stroke patients were dispensed an anticoagulant or antiplatelet agent. However, there was under-use of guideline-recommended antihypertensives (40%), statins (80%), and anticoagulants in those with AF (77%), particularly in older people or those with more comorbidities [9]. There was no major change in the proportion of people treated during the COVID-19 period. Home-time in the first 6 months, an indicator of post-stroke disability, was lower in older people, those with a more severe stroke, or more comorbidities, with modest differences by region, sex and ethnicity. After the COVID-19 period, people spent more time at home.

Secondary prevention is key to reducing stroke incidence. Whilst population-wide prescriptions of antihypertensive and lipid-lowering drugs fell during the COVID-19 pandemic[8], in stroke patients we found no major difference in dispensed medicines by year or by patient characteristics. This indicates a longer-term and systemic problem not due to the pandemic [10]. The evidence is clear that targeting a systolic blood pressure of ≤125mmHg [11] in all patients with stroke (whether ischaemic or haemorrhagic) reduces recurrent stroke, and that most stroke patients have a blood pressure over the threshold for hypertension diagnosis at presentation[12]. It is most likely that antihypertensives have substantially lower rates of dispensing - particularly in older people who stand to gain the most - and that many more patients could benefit. While these findings suggest a potential gap in secondary prevention, the use of dispensing data alone cannot account for clinical contraindications, patient preferences, or end-of-life care goals that may appropriately preclude certain medications from being prescribed. Consequently, the observed lower rates of dispensing may reflect, in part, appropriate clinical decision-making that is not captured in routine EHR records.

Linked data for measuring quality of care are essential for a learning health system[13]. Here, we developed a large, linked dataset with ascertainment of quality beyond the hospital system to primary care and pharmacy. However, this work is unlikely to be replicable routinely beyond the pandemic unless the recommendations of the Sudlow review[14] - to allow national linkage of primary care data - are implemented. No timeline is available for providing access to such data, meaning that patients with stroke who do not become inpatients (for example discharged from emergency departments, or looked after in an ambulatory TIA/stroke clinic, or diagnosed in primary care) are missing from national measurement of quality. This is particularly important for pandemic preparedness as our work shows that using a single source for ascertaining stroke, e.g. only hospitalisation data, would underestimate the number of people with stroke in England.

Our analysis has a number of strengths. First, it ascertains stroke patients in the whole English population who are registered in primary care (almost the entire population[15]). Second, we ascertained stroke from multiple overlapping sources, including the national quality register of hospitalised stroke, hospital administrative data and primary care. The date of stroke was largely consistent between sources. Third, we used previously developed and validated code lists to identify strokes. Fourth, we were able to ascertain all relevant secondary prevention medicines dispensed to patients, where a submission had been made for reimbursement from NHS England, most prescriptions in the population and a more accurate proxy of compliance than prescribing data.

Our analysis has several weaknesses. First, we were unable to check the coded diagnosis of stroke against written records. The diagnosis of stroke in health systems data in the UK[16], compared with manual review of records, has good positive predictive value (PPV), when in the primary position in hospital records (94%), is reasonable in primary care (80%), although is less good in death records (57%), and on average is less good for haemorrhagic (52%) than ischaemic stroke (83%). This is important, because risk factors and case fatality differ by subtype. Accurate clinical coding is important for all health systems, and methods should be improved with greater use of clinicians or automated coding systems to aid clinical coders [17]. Second, we did not include recurrent stroke, because differentiating further consultations with the first stroke is difficult to separate from recurrent stroke in records. Third, we examined NHS dispensed rather than prescribed medicines. This will miss medicines that were prescribed but not dispensed, which in some cases is a substantial proportion[18]. A further limitation is that our data lacks granular information on clinical intent, medication intolerance, and contraindications. We cannot distinguish between a failure to prescribe and a deliberate clinical decision based on patient frailty or preferences. It will also miss prescriptions from the private sector which are not collected through NHS reimbursement schemes. Fourth, in this manuscript, we did not examine in detail differences by ethnicity or geography and so may have missed communities who were particularly disadvantaged. Fifth, whilst home-time does correlate with disability and symptoms in this analysis and others, it is influenced by other factors such as family needs and does not directly measure impairment [19–21]. Sixth, our estimates of stroke incidence were high, compared with population-based incidence studies with bespoke measurement of stroke. This may be because there is overuse of stroke codes in primary care data, or that differentiating first-ever from recurrent stroke is difficult. This emphasises the need for improved clinical coding throughout healthcare pathways. Seventh, we used data provided by SSNAP under the Notice under Regulation 3(4) of the Health Service (Control of Patient Information) Regulations 200, that under provisioned records because of truncated monthly uploads to the SDE. Planning mechanisms for complete flow of data would prepare for the next health emergency.

Our analysis has several implications for health policy makers and stroke clinicians. Secondary prevention should be a priority for healthcare quality improvement for patients with stroke, and investigation of the clinical, system, and patient related factors leading to underuse should be research priorities. Despite the disruption caused by the COVID-19 pandemic on secondary prevention, we have shown here that there is longer term underuse that must be addressed. Home-time is an indicator of disability which could be added to the SSNAP and the stroke audit in Scotland for all patients, if it is interpreted carefully. Lastly, the use of linked health data - with an awareness of its limitations - is a promising avenue for audits of care for people with stroke and other conditions, to maximise ascertainment, streamline efficiency of data collection and enhance utility.

## Acknowledgements

This study made use of de-identified data held in NHS England’s SDE service for England and made available via the BHF Data Science Centre’s CVD-COVID-UK/COVID-IMPACT consortium. This work used data provided by patients and collected by the NHS as part of their care and support. We would also like to acknowledge all data providers who make health relevant data available for research.

## Author contributions

S.D.and W.W. conceived the research. S. D., J.F., J.N., S.L., J.M. and W.W. contributed to the design and planning of the research. J.F carried out the analysis. S. D., J.F., J.N., S.L., R.L., M.K., J.M., A.W. and W.W contributed to the interpretation of the results. S. D., J.F., J.N., S.L., J.M., and W.W. wrote the first draft of the manuscript. All authors reviewed and commented on the manuscript.

## References

1. Primary S. Sentinel Stroke National Audit Programme (SSNAP). London: Royal College of Physcians. 2014. Available: https://www.hqip.org.uk/wp-content/uploads/2019/06/Ref-142-SSNAP-Annual-Report-FINAL.pdf

2. Herrett E, Shah AD, Boggon R, Denaxas S, Smeeth L, van Staa T, et al. Completeness and diagnostic validity of recording acute myocardial infarction events in primary care, hospital care, disease registry, and national mortality records: cohort study. BMJ. 2013;346: f2350.

3. Wood A, Denholm R, Hollings S, Cooper J, Ip S, Walker V, et al. Linked electronic health records for research on a nationwide cohort of more than 54 million people in England: data resource. BMJ. 2021;373: n826.

4. Dispensing data. [cited 12 Mar 2025]. Available: https://www.nhsbsa.nhs.uk/prescription-data/dispensing-data

5. Campbell A. Quality of mortality data during the coronavirus pandemic, England and Wales - Office for National Statistics. Office for National Statistics; 3 Dec 2020 [cited 26 Jan 2022]. Available: https://www.ons.gov.uk/peoplepopulationandcommunity/birthsdeathsandmarriages/deaths/articles/qualityofmortalitydataduringthecoronaviruspandemicenglandandwales/2020

6. List of ethnic groups. [cited 12 Mar 2025]. Available: https://www.ethnicity-facts-figures.service.gov.uk/style-guide/ethnic-groups/

7. Thygesen JH, Tomlinson C, Hollings S, Mizani MA, Handy A, Akbari A, et al. COVID-19 trajectories among 57 million adults in England: a cohort study using electronic health records. Lancet Digit Health. 2022;4: e542–e557.

8. Dale CE, Takhar R, Carragher R, Katsoulis M, Torabi F, Duffield S, et al. The impact of the COVID-19 pandemic on cardiovascular disease prevention and management. Nat Med. 2023;29: 219–225.

9. National Clinical Guideline for Stroke. In: National Clinical Guideline for Stroke [Internet]. 5 Oct 2022 [cited 12 Mar 2025]. Available: https://www.strokeguideline.org/

10. Whitty CJM, Smith G, McBride M, Atherton F, Powis SH, Stokes-Lampard H. Restoring and extending secondary prevention. BMJ. 2023;380: 201.

11. Long-term management and secondary prevention. In: National Clinical Guideline for Stroke [Internet]. 23 Jan 2023 [cited 12 Mar 2025]. Available: https://www.strokeguideline.org/chapter/long-term-management-and-secondary-prevention/

12. Fischer U, Cooney MT, Bull LM, Silver LE, Chalmers J, Anderson CS, et al. Acute post-stroke blood pressure relative to premorbid levels in intracerebral haemorrhage versus major ischaemic stroke: a population-based study. Lancet Neurol. 2014;13: 374–384.

13. Cadilhac DA, Bravata DM, Bettger JP, Mikulik R, Norrving B, Uvere EO, et al. Stroke learning health systems: A topical narrative review with case examples. Stroke. 2023;54: 1148–1159.

14. Sudlow C. The Sudlow Review. In: HDR UK [Internet]. 31 May 2023 [cited 12 Mar 2025]. Available: https://www.hdruk.ac.uk/helping-with-health-data/the-sudlow-review/

15. Patients Registered at a GP Practice, February 2025. In: NHS England Digital [Internet]. [cited 12 Mar 2025]. Available: https://digital.nhs.uk/data-and-information/publications/statistical/patients-registered-at-a-gp-practice/february-2025

16. Rannikmäe K, Ngoh K, Bush K, Al-Shahi Salman R, Doubal F, Flaig R, et al. Accuracy of identifying incident stroke cases from linked health care data in UK Biobank. Neurology. 2020;95: e697–e707.

17. Dong H, Falis M, Whiteley W, Alex B, Matterson J, Ji S, et al. Automated clinical coding: what, why, and where we are? NPJ Digit Med. 2022;5: 159.

18. Gardner TL, Dovey SM, Tilyard MW, Gurr E. Differences between prescribed and dispensed medications. N Z Med J. 1996;109: 69–72.

19. McDermid I, Barber M, Dennis M, Langhorne P, Macleod MJ, McAlpine CH, et al. Home-time is a feasible and valid stroke outcome measure in national datasets. Stroke. 2019;50: 1282–1285.

20. Shen E, Rozema EJ, Haupt EC, Henry M, Scholle SH, Wang SE, et al. Assessing the concurrent validity of days alive and at home metric. J Am Geriatr Soc. 2022;70: 2630–2637.

21. Sung S-F, Su C-C, Hsieh C-Y, Cheng C-L, Chen C-H, Lin H-J, et al. Home-time as a surrogate measure for functional outcome after stroke: A validation study. Clin Epidemiol. 2020;12: 617–624.

