## Supplementary material for "Measurement of quality of stroke care with national electronic health records: a prospective cohort study during and after the COVID-19 pandemic"

### Tables

Table S1 - Age- and sex-standardised incidence rates of stroke (standardised to the European Standard Population) across calendar years 2020 to 2023 for individuals aged 18 years and older. Values are presented as rates per 100,000 population with 95% confidence intervals.

|  |  | **2020** | **2021** | **2022** | **2023** |
| --- | --- | --- | --- | --- | --- |
| **Overall** | Overall | 227.6 (226.1, 229.0) | 244.0 (242.6, 245.5) | 242.2 (240.8, 243.7) | 244.8 (243.4, 246.3) |
| **Stroke type** | Ischaemic | 157.6 (156.4, 158.8) | 170.1 (168.9, 171.4) | 168.9 (167.6, 170.1) | 171.9 (170.7, 173.1) |
|  | Haemorrhagic | 29.5 (29.0, 30.0) | 31.7 (31.1, 32.2) | 31.7 (31.2, 32.2) | 31.9 (31.4, 32.4) |
|  | Unknown | 40.5 (39.9, 41.1) | 42.3 (41.6, 42.9) | 41.7 (41.1, 42.3) | 41.0 (40.4, 41.6) |
| **Sex** | Female | 196.8 (195.0, 198.6) | 210.1 (208.2, 211.9) | 208.8 (207.0, 210.6) | 209.4 (207.6, 211.2) |
|  | Male | 258.4 (256.1, 260.6) | 278.0 (275.7, 280.3) | 275.7 (273.4, 278.0) | 280.2 (277.9, 282.5) |
| **Age group (years)** | 18-59 | 60.2 (59.3, 61.1) | 66.5 (65.6, 67.4) | 65.2 (64.3, 66.1) | 66.7 (65.8, 67.6) |
|  | 60-69 | 287.1 (282.8, 291.5) | 313.3 (308.9, 317.8) | 310.3 (306.0, 314.8) | 311.9 (307.6, 316.3) |
|  | 70-79 | 554.1 (547.4, 560.9) | 594.9 (588.0, 601.8) | 603.4 (596.5, 610.3) | 607.0 (600.2, 614.0) |
|  | 80-89 | 1188.8 (1174.6, 1203.2) | 1257.8 (1243.2, 1272.6) | 1232.6 (1218.3, 1247.1) | 1248.1 (1233.8, 1262.5) |
|  | 90+ | 1980.9 (1939.8, 2022.7) | 2013.5 (1972.2, 2055.4) | 2002.1 (1961.6, 2043.2) | 2015.6 (1974.9, 2056.9) |
| **Region** | East Midlands | 231.7 (226.9, 236.6) | 252.2 (247.2, 257.2) | 244.5 (239.6, 249.4) | 249.4 (244.6, 254.4) |
|  | East of England | 209.3 (205.3, 213.3) | 223.1 (219.0, 227.3) | 219.5 (215.5, 223.6) | 223.6 (219.6, 227.7) |
|  | London | 224.0 (219.7, 228.3) | 243.0 (238.5, 247.5) | 241.5 (237.1, 245.9) | 247.0 (242.6, 251.5) |
|  | North East | 248.6 (242.0, 255.4) | 280.0 (272.9, 287.1) | 283.8 (276.7, 290.9) | 277.5 (270.6, 284.6) |
|  | North West | 244.2 (240.1, 248.3) | 259.4 (255.2, 263.6) | 258.6 (254.4, 262.8) | 260.4 (256.2, 264.5) |
|  | South East | 219.7 (216.3, 223.1) | 233.4 (230.0, 236.9) | 227.1 (223.7, 230.5) | 227.1 (223.7, 230.5) |
|  | South West | 224.6 (220.5, 228.8) | 236.5 (232.3, 240.8) | 238.8 (234.5, 243.0) | 246.6 (242.3, 250.9) |
|  | West Midlands | 219.6 (215.3, 223.9) | 231.3 (227.0, 235.8) | 233.7 (229.3, 238.1) | 228.9 (224.6, 233.3) |
|  | Yorkshire and The Humber | 244.7 (240.0, 249.5) | 265.6 (260.7, 270.5) | 268.3 (263.4, 273.3) | 275.2 (270.3, 280.2) |
| **Data source membership** | GDPPR | 164.8 (163.6, 166.0) | 175.3 (174.0, 176.5) | 173.0 (171.8, 174.3) | 172.6 (171.3, 173.8) |
|  | HES-APC | 149.9 (148.8, 151.1) | 161.6 (160.4, 162.8) | 157.7 (156.5, 158.9) | 160.5 (159.4, 161.7) |
|  | SSNAP | 125.9 (124.9, 127.0) | 125.2 (124.1, 126.3) | 117.3 (116.3, 118.4) | 128.2 (127.2, 129.3) |
|  | ONS Deaths | 34.6 (34.1, 35.2) | 35.3 (34.7, 35.8) | 32.3 (31.8, 32.8) | 31.1 (30.6, 31.7) |
|  | GDPPR or HES-APC | 209.1 (207.8, 210.5) | 225.7 (224.3, 227.1) | 226.0 (224.6, 227.4) | 228.7 (227.3, 230.1) |
|  | GDPPR or SSNAP | 199.2 (197.8, 200.5) | 210.4 (209.0, 211.8) | 208.9 (207.6, 210.3) | 213.8 (212.5, 215.2) |
|  | HES-APC or SSNAP | 159.9 (158.7, 161.1) | 170.8 (169.6, 172.1) | 166.1 (164.9, 167.3) | 169.6 (168.4, 170.8) |
|  | GDPPR or HES-APC or SSNAP or ONS Deaths | 227.6 (226.1, 229.0) | 244.0 (242.6, 245.5) | 242.2 (240.8, 243.7) | 244.8 (243.4, 246.3) |

Table S2.a - Contingency table of stroke records by data source and stroke type in individuals with non-fatal stroke (n=358,025) identified in GDPPR, HES-APC or SSNAP. Values represent the number of individuals, with column percentages in parentheses. NOS: not otherwise specified stroke type.

| **Data source & stroke type** | **SSNAP** | | | **HES-APC** | | | **GDPPR** | | | **All non-fatal strokes (n=358025)** |
| --- | --- | --- | --- | --- | --- | --- | --- | --- | --- | --- |
|  | **Ischaemic stroke (n=173965)** | **Haemorrhagic stroke (n=19210)** | **Missing stroke type**  **(n=725)** | **Ischaemic stroke**  **(n=207990)** | **Haemorrhagic stroke**  **(n=26555)** | **NOS stroke**  **(n=7815)** | **Ischaemic stroke**  **(n=126040)** | **Haemorrhagic stroke**  **(n=22860)** | **NOS stroke**  **(n=162880)** |  |
| **SSNAP** |  |  |  |  |  |  |  |  |  |  |
| Ischaemic | 173965 (100.0) | 0 (0.0) | 105 (14.5) | 157470 (75.7) | 1385 (5.2) | 4405 (56.4) | 67065 (53.2) | 1510 (6.6) | 79415 (48.8) | 173965 (48.6) |
| Haemorrhagic | 0 (0.0) | 19210 (100.0) | 0 (0.0) | 1275 (0.6) | 15945 (60.0) | 40 (0.5) | 760 (0.6) | 9495 (41.5) | 5025 (3.1) | 19210 (5.4) |
| Missing type | 105 (0.1) | 0 (0.0) | 725 (100.0) | 560 (0.3) | 55 (0.2) | 20 (0.3) | 295 (0.2) | 35 (0.2) | 280 (0.2) | 725 (0.2) |
| No record | 0 (0.0) | 0 (0.0) | 0 (0.0) | 48785 (23.5) | 9180 (34.6) | 3350 (42.9) | 57975 (46.0) | 11830 (51.7) | 78195 (48.0) | 164245 (45.9) |
| **HES-APC** |  |  |  |  |  |  |  |  |  |  |
| Ischaemic | 157470 (90.5) | 1275 (6.6) | 560 (77.2) | 207990 (100.0) | 245 (0.9) | 1080 (13.8) | 79795 (63.3) | 1690 (7.4) | 92860 (57.0) | 207990 (58.1) |
| Haemorrhagic | 1385 (0.8) | 15945 (83.0) | 55 (7.6) | 245 (0.1) | 26555 (100.0) | 15 (0.2) | 830 (0.7) | 12715 (55.6) | 6005 (3.7) | 26555 (7.4) |
| NOS | 4405 (2.5) | 40 (0.2) | 20 (2.8) | 1080 (0.5) | 15 (0.1) | 7815 (100.0) | 1430 (1.1) | 45 (0.2) | 3605 (2.2) | 7815 (2.2) |
| No record | 11625 (6.7) | 2060 (10.7) | 90 (12.4) | 0 (0.0) | 0 (0.0) | 0 (0.0) | 44365 (35.2) | 8470 (37.1) | 61015 (37.5) | 117000 (32.7) |
| **GDPPR** |  |  |  |  |  |  |  |  |  |  |
| Ischaemic | 67065 (38.6) | 760 (4.0) | 295 (40.7) | 79795 (38.4) | 830 (3.1) | 1430 (18.3) | 126040 (100.0) | 815 (3.6) | 18720 (11.5) | 126040 (35.2) |
| Haemorrhagic | 1510 (0.9) | 9495 (49.4) | 35 (4.8) | 1690 (0.8) | 12715 (47.9) | 45 (0.6) | 815 (0.6) | 22860 (100.0) | 2695 (1.7) | 22860 (6.4) |
| NOS | 79415 (45.6) | 5025 (26.2) | 280 (38.6) | 92860 (44.6) | 6005 (22.6) | 3605 (46.1) | 18720 (14.9) | 2695 (11.8) | 162880 (100.0) | 162880 (45.5) |
| No record | 39565 (22.7) | 5830 (30.3) | 180 (24.8) | 49580 (23.8) | 9270 (34.9) | 3065 (39.2) | 0 (0.0) | 0 (0.0) | 0 (0.0) | 68315 (19.1) |

Table S2.b - Contingency table of stroke records by data source and stroke type in individuals with a fatal stroke (n=67,650) identified in GDPPR, HES-APC, SSNAP or ONS Deaths. Values represent the number of individuals, with column percentages in parentheses. NOS: not otherwise specified stroke type.

| **Data source & stroke type** | **SSANP** | | | **HES-ACP** | | | **GDPPR** | | | **ONS Deaths** | | | **All fatal strokes**  **(n=67650)** |
| --- | --- | --- | --- | --- | --- | --- | --- | --- | --- | --- | --- | --- | --- |
|  | **Ischaemic stroke (n=19170)** | **Haemorrhagic stroke**  **(n=7440)** | **Missing stroke type**  **(n=100)** | **Ischaemic stroke**  **(n=25355)** | **Haemorrhagic stroke**  **(n=12580)** | **NOS stroke**  **(n=1045)** | **Ischaemic stroke**  **(n=3055)** | **Haemorrhagic stroke**  **(n=3635)** | **NOS stroke**  **(n=8420)** | **Ischaemic stroke**  **(n=14415)** | **Haemorrhagic stroke**  **(n=15615)** | **NOS stroke**  **(n=31760)** |  |
| **SSNAP** |  |  |  |  |  |  |  |  |  |  |  |  |  |
| Ischaemic | 19170 (100.0) | 0 (0.0) | 15 (15.0) | 17035 (67.2) | 365 (2.9) | 385 (36.8) | 1770 (57.9) | 90 (2.5) | 3635 (43.2) | 7130 (49.5) | 710 (4.5) | 10975 (34.6) | 19170 (28.3) |
| Haemorrhagic | 0 (0.0) | 7440 (100.0) | 0 (0.0) | 250 (1.0) | 6180 (49.1) | 0 (0.0) | 40 (1.3) | 1225 (33.7) | 625 (7.4) | 90 (0.6) | 5235 (33.5) | 560 (1.8) | 7440 (11.0) |
| Missing type | 15 (0.1) | 0 (0.0) | 100 (100.0) | 65 (0.3) | 10 (0.1) | 0 (0.0) | 10 (0.3) | 0 (0.0) | 15 (0.2) | 25 (0.2) | 10 (0.1) | 55 (0.2) | 100 (0.1) |
| No stroke record | 0 (0.0) | 0 (0.0) | 0 (0.0) | 8015 (31.6) | 6025 (47.9) | 645 (61.7) | 1235 (40.4) | 2320 (63.8) | 4145 (49.2) | 7175 (49.8) | 9660 (61.9) | 20175 (63.5) | 40960 (60.5) |
| **HES-APC** |  |  |  |  |  |  |  |  |  |  |  |  |  |
| Ischaemic | 17035 (88.9) | 250 (3.4) | 65 (65.0) | 25355 (100.0) | 75 (0.6) | 95 (9.1) | 2270 (74.3) | 130 (3.6) | 4525 (53.7) | 9640 (66.9) | 845 (5.4) | 14210 (44.7) | 25355 (37.5) |
| Haemorrhagic | 365 (1.9) | 6180 (83.1) | 10 (10.0) | 75 (0.3) | 12580 (100.0) | 0 (0.0) | 50 (1.6) | 2105 (57.9) | 760 (9.0) | 195 (1.4) | 8825 (56.5) | 785 (2.5) | 12580 (18.6) |
| NOS | 385 (2.0) | 0 (0.0) | 0 (0.0) | 95 (0.4) | 0 (0.0) | 1045 (100.0) | 35 (1.1) | 0 (0.0) | 150 (1.8) | 125 (0.9) | 25 (0.2) | 720 (2.3) | 1045 (1.5) |
| No stroke record | 1490 (7.8) | 1010 (13.6) | 20 (20.0) | 0 (0.0) | 0 (0.0) | 0 (0.0) | 710 (23.2) | 1395 (38.4) | 3020 (35.9) | 4510 (31.3) | 5960 (38.2) | 16145 (50.8) | 28850 (42.6) |
| **GDPPR** |  |  |  |  |  |  |  |  |  |  |  |  |  |
| Ischaemic | 1770 (9.2) | 40 (0.5) | 10 (10.0) | 2270 (9.0) | 50 (0.4) | 35 (3.3) | 3055 (100.0) | 35 (1.0) | 405 (4.8) | 1115 (7.7) | 100 (0.6) | 1405 (4.4) | 3055 (4.5) |
| Haemorrhagic | 90 (0.5) | 1225 (16.5) | 0 (0.0) | 130 (0.5) | 2105 (16.7) | 0 (0.0) | 35 (1.1) | 3635 (100.0) | 165 (2.0) | 80 (0.6) | 2245 (14.4) | 190 (0.6) | 3635 (5.4) |
| NOS | 3635 (19.0) | 625 (8.4) | 15 (15.0) | 4525 (17.8) | 760 (6.0) | 150 (14.4) | 405 (13.3) | 165 (4.5) | 8420 (100.0) | 1700 (11.8) | 815 (5.2) | 4670 (14.7) | 8420 (12.4) |
| No stroke record | 13995 (73.0) | 5645 (75.9) | 80 (80.0) | 18815 (74.2) | 9785 (77.8) | 865 (82.8) | 0 (0.0) | 0 (0.0) | 0 (0.0) | 11705 (81.2) | 12595 (80.7) | 25750 (81.1) | 53145 (78.6) |
| **ONS Deaths** |  |  |  |  |  |  |  |  |  |  |  |  |  |
| Ischaemic | 7130 (37.2) | 90 (1.2) | 25 (25.0) | 9640 (38.0) | 195 (1.6) | 125 (12.0) | 1115 (36.5) | 80 (2.2) | 1700 (20.2) | 14415 (100.0) | 465 (3.0) | 1215 (3.8) | 14415 (21.3) |
| Haemorrhagic | 710 (3.7) | 5235 (70.4) | 10 (10.0) | 845 (3.3) | 8825 (70.2) | 25 (2.4) | 100 (3.3) | 2245 (61.8) | 815 (9.7) | 465 (3.2) | 15615 (100.0) | 665 (2.1) | 15615 (23.1) |
| NOS | 10975 (57.3) | 560 (7.5) | 55 (55.0) | 14210 (56.0) | 785 (6.2) | 720 (68.9) | 1405 (46.0) | 190 (5.2) | 4670 (55.5) | 1215 (8.4) | 665 (4.3) | 31760 (100.0) | 31760 (46.9) |
| No stroke record | 1445 (7.5) | 1740 (23.4) | 15 (15.0) | 2085 (8.2) | 3120 (24.8) | 185 (17.7) | 585 (19.1) | 1205 (33.1) | 1530 (18.2) | 0 (0.0) | 0 (0.0) | 0 (0.0) | 8185 (12.1) |

Table S3 - Contingency table of stroke records by data source, stroke type and electronic health record code in strokes identified in GDPPR, HES-APC, SSNAP or ONS Deaths. Values represent the number of individuals, with column percentages in parentheses.

[Provided as .csv file: R04-stroke_phenotype_breakdown / contingency_tbl_code_cohort.csv]

Table S4.a - Cause-specific multivariable Cox proportional hazard regression for medications dispensed in primary care for non-fatal strokes (n = 348,150).

|  |  | Cause-specific hazard ratio ([95% confidence interval], p-value) | | | | |
| --- | --- | --- | --- | --- | --- | --- |
|  |  | Anticoagulants or Antiplatelets | Anticoagulants | Antiplatelets | Antihypertensives | Lipid lowering drugs |
| Year of stroke | 2020 | (Ref.) | (Ref.) | (Ref.) | (Ref.) | (Ref.) |
|  | 2021 | 1.01 ([1.00, 1.02], p=0.021) | 1.03 ([1.01, 1.04], p=0.010) | 1.01 ([0.99, 1.02], p=0.374) | 1.04 ([1.03, 1.06], p<0.001) | 1.04 ([1.03, 1.05], p<0.001) |
|  | 2022 | 1.01 ([1.00, 1.02], p=0.035) | 1.02 ([1.00, 1.04], p=0.016) | 1.01 ([1.00, 1.02], p=0.221) | 1.05 ([1.03, 1.07], p<0.001) | 1.06 ([1.05, 1.07], p<0.001) |
|  | 2023 | 1.00 ([0.99, 1.01], p=0.455) | 1.02 ([1.00, 1.04], p=0.031) | 1.00 ([0.99, 1.01], p=0.893) | 1.06 ([1.05, 1.08], p<0.001) | 1.09 ([1.08, 1.10], p<0.001) |
| Age group (years) | 18-59 | (Ref.) | (Ref.) | (Ref.) | (Ref.) | (Ref.) |
|  | 60-69 | 1.22 ([1.21, 1.23], p<0.001) | 1.14 ([1.11, 1.17], p<0.001) | 1.18 ([1.17, 1.20], p<0.001) | 0.97 ([0.96, 0.99], p=0.002) | 1.21 ([1.20, 1.23], p<0.001) |
|  | 70-79 | 1.21 ([1.19, 1.22], p<0.001) | 1.25 ([1.22, 1.28], p<0.001) | 1.13 ([1.11, 1.14], p<0.001) | 0.90 ([0.88, 0.91], p<0.001) | 1.17 ([1.16, 1.18], p<0.001) |
|  | 80-89 | 1.21 ([1.20, 1.23], p<0.001) | 1.24 ([1.21, 1.27], p<0.001) | 1.09 ([1.07, 1.10], p<0.001) | 0.76 ([0.74, 0.77], p<0.001) | 1.04 ([1.02, 1.05], p<0.001) |
|  | 90+ | 1.17 ([1.15, 1.19], p<0.001) | 1.09 ([1.05, 1.12], p<0.001) | 1.06 ([1.04, 1.09], p<0.001) | 0.56 ([0.55, 0.58], p<0.001) | 0.62 ([0.61, 0.64], p<0.001) |
| Sex | Female | (Ref.) | (Ref.) | (Ref.) | (Ref.) | (Ref.) |
|  | Male | 1.01 ([1.00, 1.02], p=0.053) | 0.99 ([0.98, 1.01], p=0.463) | 1.03 ([1.02, 1.04], p<0.001) | 1.08 ([1.07, 1.09], p<0.001) | 1.07 ([1.06, 1.08], p<0.001) |
| IMD 2019 quintiles | 1 (most deprived) | (Ref.) | (Ref.) | (Ref.) | (Ref.) | (Ref.) |
|  | 2 | 0.98 ([0.97, 0.99], p<0.001) | 1.00 ([0.98, 1.02], p=0.859) | 0.98 ([0.96, 0.99], p=0.001) | 1.00 ([0.99, 1.02], p=0.693) | 0.98 ([0.97, 0.99], p=0.001) |
|  | 3 | 0.97 ([0.96, 0.98], p<0.001) | 1.01 ([0.99, 1.03], p=0.442) | 0.97 ([0.95, 0.98], p<0.001) | 1.01 ([1.00, 1.03], p=0.076) | 0.97 ([0.96, 0.98], p<0.001) |
|  | 4 | 0.95 ([0.94, 0.97], p<0.001) | 1.00 ([0.98, 1.02], p=0.775) | 0.95 ([0.94, 0.96], p<0.001) | 1.02 ([1.01, 1.04], p=0.003) | 0.96 ([0.95, 0.97], p<0.001) |
|  | 5 (least deprived) | 0.94 ([0.93, 0.95], p<0.001) | 1.01 ([0.99, 1.03], p=0.512) | 0.93 ([0.92, 0.95], p<0.001) | 1.01 ([0.99, 1.03], p=0.199) | 0.96 ([0.95, 0.97], p<0.001) |
| Ethnicity | White | (Ref.) | (Ref.) | (Ref.) | (Ref.) | (Ref.) |
|  | Asian | 0.89 ([0.87, 0.90], p<0.001) | 0.79 ([0.76, 0.82], p<0.001) | 0.92 ([0.90, 0.93], p<0.001) | 1.10 ([1.07, 1.12], p<0.001) | 0.95 ([0.94, 0.97], p<0.001) |
|  | Black | 0.89 ([0.87, 0.91], p<0.001) | 0.93 ([0.88, 0.97], p=0.002) | 0.90 ([0.88, 0.93], p<0.001) | 1.16 ([1.13, 1.20], p<0.001) | 0.91 ([0.89, 0.94], p<0.001) |
|  | Mixed | 0.86 ([0.82, 0.89], p<0.001) | 0.88 ([0.81, 0.96], p=0.005) | 0.87 ([0.83, 0.92], p<0.001) | 1.02 ([0.97, 1.08], p=0.450) | 0.87 ([0.83, 0.91], p<0.001) |
|  | Other | 0.86 ([0.83, 0.89], p<0.001) | 0.91 ([0.84, 0.98], p=0.009) | 0.88 ([0.84, 0.91], p<0.001) | 1.01 ([0.97, 1.07], p=0.552) | 0.91 ([0.87, 0.94], p<0.001) |
|  | Unknown | 0.91 ([0.84, 0.98], p=0.008) | 1.05 ([0.91, 1.22], p=0.510) | 0.89 ([0.82, 0.96], p=0.003) | 1.15 ([1.04, 1.27], p=0.006) | 0.89 ([0.82, 0.96], p=0.002) |
| Region | East Midlands | (Ref.) | (Ref.) | (Ref.) | (Ref.) | (Ref.) |
|  | East of England | 0.93 ([0.92, 0.95], p<0.001) | 1.04 ([1.01, 1.08], p=0.004) | 0.94 ([0.92, 0.96], p<0.001) | 0.93 ([0.91, 0.95], p<0.001) | 0.94 ([0.92, 0.96], p<0.001) |
|  | London | 0.80 ([0.79, 0.82], p<0.001) | 1.00 ([0.97, 1.03], p=0.921) | 0.82 ([0.80, 0.83], p<0.001) | 0.86 ([0.84, 0.88], p<0.001) | 0.84 ([0.82, 0.85], p<0.001) |
|  | North East | 1.00 ([0.98, 1.02], p=0.708) | 0.91 ([0.88, 0.95], p<0.001) | 1.02 ([1.00, 1.05], p=0.030) | 1.00 ([0.97, 1.02], p=0.807) | 1.00 ([0.98, 1.02], p=0.635) |
|  | North West | 0.98 ([0.96, 0.99], p=0.006) | 0.98 ([0.96, 1.01], p=0.295) | 0.99 ([0.97, 1.00], p=0.126) | 0.97 ([0.95, 0.99], p=0.001) | 0.95 ([0.94, 0.97], p<0.001) |
|  | South East | 0.88 ([0.87, 0.89], p<0.001) | 0.98 ([0.95, 1.01], p=0.173) | 0.90 ([0.89, 0.92], p<0.001) | 0.88 ([0.86, 0.90], p<0.001) | 0.87 ([0.86, 0.88], p<0.001) |
|  | South West | 0.90 ([0.89, 0.92], p<0.001) | 0.95 ([0.92, 0.98], p=0.001) | 0.93 ([0.91, 0.95], p<0.001) | 0.88 ([0.86, 0.90], p<0.001) | 0.86 ([0.85, 0.88], p<0.001) |
|  | West Midlands | 0.91 ([0.89, 0.92], p<0.001) | 0.97 ([0.94, 1.00], p=0.026) | 0.93 ([0.91, 0.95], p<0.001) | 0.90 ([0.88, 0.92], p<0.001) | 0.91 ([0.89, 0.92], p<0.001) |
|  | Yorkshire and The Humber | 1.02 ([1.00, 1.04], p=0.017) | 1.01 ([0.98, 1.04], p=0.667) | 1.02 ([1.00, 1.04], p=0.021) | 0.98 ([0.96, 1.01], p=0.137) | 1.03 ([1.01, 1.05], p=0.001) |
| Charlson comorbidity index | 0 | (Ref.) | (Ref.) | (Ref.) | (Ref.) | (Ref.) |
|  | 1-2 | 1.01 ([1.00, 1.03], p=0.012) | 0.94 ([0.92, 0.96], p<0.001) | 1.08 ([1.06, 1.09], p<0.001) | 1.01 ([1.00, 1.03], p=0.090) | 1.00 ([0.99, 1.01], p=0.881) |
|  | 3-4 | 0.99 ([0.98, 1.00], p=0.105) | 0.93 ([0.91, 0.95], p<0.001) | 1.07 ([1.06, 1.09], p<0.001) | 0.96 ([0.94, 0.97], p<0.001) | 0.94 ([0.93, 0.95], p<0.001) |
|  | 5+ | 0.95 ([0.94, 0.97], p<0.001) | 0.88 ([0.85, 0.90], p<0.001) | 1.08 ([1.07, 1.10], p<0.001) | 0.84 ([0.82, 0.86], p<0.001) | 0.85 ([0.84, 0.87], p<0.001) |
| Data source combination | GDPPR only | (Ref.) | (Ref.) | (Ref.) | (Ref.) | (Ref.) |
|  | HES-APC only | 0.94 ([0.92, 0.95], p<0.001) | 1.21 ([1.18, 1.25], p<0.001) | 0.82 ([0.80, 0.84], p<0.001) | 0.93 ([0.91, 0.96], p<0.001) | 0.94 ([0.93, 0.96], p<0.001) |
|  | SSNAP only | 0.84 ([0.82, 0.87], p<0.001) | 1.14 ([1.08, 1.20], p<0.001) | 0.74 ([0.72, 0.77], p<0.001) | 0.98 ([0.95, 1.03], p=0.448) | 0.91 ([0.88, 0.94], p<0.001) |
|  | GDPPR & HES-APC | 1.37 ([1.35, 1.39], p<0.001) | 1.39 ([1.36, 1.42], p<0.001) | 1.23 ([1.21, 1.25], p<0.001) | 1.12 ([1.10, 1.14], p<0.001) | 1.32 ([1.30, 1.34], p<0.001) |
|  | GDPPR & SSNAP | 1.26 ([1.22, 1.29], p<0.001) | 1.29 ([1.22, 1.35], p<0.001) | 1.17 ([1.13, 1.21], p<0.001) | 1.21 ([1.17, 1.25], p<0.001) | 1.28 ([1.24, 1.32], p<0.001) |
|  | HES-APC & SSNAP | 1.26 ([1.25, 1.28], p<0.001) | 1.38 ([1.35, 1.42], p<0.001) | 1.10 ([1.08, 1.12], p<0.001) | 1.08 ([1.06, 1.10], p<0.001) | 1.26 ([1.24, 1.28], p<0.001) |
|  | GDPPR & HES-APC & SSNAP | 1.50 ([1.49, 1.51], p<0.001) | 1.46 ([1.43, 1.49], p<0.001) | 1.33 ([1.32, 1.34], p<0.001) | 1.21 ([1.19, 1.22], p<0.001) | 1.50 ([1.48, 1.51], p<0.001) |
| Atrial fibrillation | No | (Ref.) | (Ref.) | (Ref.) | (Ref.) | (Ref.) |
|  | Yes | 1.14 ([1.13, 1.15], p<0.001) | 6.33 ([6.21, 6.45], p<0.001) | 0.29 ([0.28, 0.29], p<0.001) | 0.92 ([0.91, 0.94], p<0.001) | 0.98 ([0.97, 0.99], p<0.001) |
| Pre-stroke anticoagulants | No | (Ref.) | (Ref.) | (Ref.) | (Ref.) | (Ref.) |
|  | Yes | 1.16 ([1.14, 1.17], p<0.001) | 3.08 ([3.02, 3.13], p<0.001) | 0.29 ([0.28, 0.29], p<0.001) | 0.95 ([0.93, 0.97], p<0.001) | 0.83 ([0.82, 0.84], p<0.001) |
| Pre-stroke antiplatelets | No | (Ref.) | (Ref.) | (Ref.) | (Ref.) | (Ref.) |
|  | Yes | 1.51 ([1.50, 1.53], p<0.001) | 0.82 ([0.81, 0.83], p<0.001) | 1.83 ([1.82, 1.85], p<0.001) | 0.99 ([0.97, 1.00], p=0.020) | 0.98 ([0.97, 0.99], p<0.001) |
| Pre-stroke antihypertensives | No | (Ref.) | (Ref.) | (Ref.) | (Ref.) | (Ref.) |
|  | Yes | 1.05 ([1.04, 1.06], p<0.001) | 1.03 ([1.02, 1.05], p<0.001) | 1.04 ([1.03, 1.05], p<0.001) | 10.41 ([10.28, 10.55], p<0.001) | 1.03 ([1.02, 1.04], p<0.001) |
| Pre-stroke lipid lowering drugs | No | (Ref.) | (Ref.) | (Ref.) | (Ref.) | (Ref.) |
|  | Yes | 1.09 ([1.08, 1.10], p<0.001) | 1.01 ([0.99, 1.02], p=0.354) | 1.06 ([1.04, 1.07], p<0.001) | 0.98 ([0.97, 1.00], p=0.006) | 2.45 ([2.43, 2.48], p<0.001) |

Table S4.b - Cause-specific multivariable Cox proportional hazard regression for medications dispensed in primary care for non-fatal ischaemic strokes or strokes of unknown type (n = 313,715).

|  |  | Cause-specific hazard ratio ([95% confidence interval], p-value) | | | | |
| --- | --- | --- | --- | --- | --- | --- |
|  |  | Anticoagulants or Antiplatelets | Anticoagulants | Antiplatelets | Antihypertensives | Lipid lowering drugs |
| Year of stroke | 2020 | (Ref.) | (Ref.) | (Ref.) | (Ref.) | (Ref.) |
|  | 2021 | 1.01 ([1.00, 1.02], p=0.032) | 1.02 ([1.00, 1.04], p=0.076) | 1.01 ([0.99, 1.02], p=0.346) | 1.04 ([1.02, 1.05], p<0.001) | 1.04 ([1.03, 1.05], p<0.001) |
|  | 2022 | 1.01 ([1.00, 1.02], p=0.030) | 1.02 ([1.00, 1.04], p=0.020) | 1.01 ([0.99, 1.02], p=0.262) | 1.04 ([1.03, 1.06], p<0.001) | 1.05 ([1.04, 1.07], p<0.001) |
|  | 2023 | 1.00 ([0.99, 1.01], p=0.549) | 1.01 ([0.99, 1.03], p=0.153) | 1.00 ([0.99, 1.01], p=0.992) | 1.06 ([1.04, 1.07], p<0.001) | 1.08 ([1.07, 1.10], p<0.001) |
| Age group (years) | 18-59 | (Ref.) | (Ref.) | (Ref.) | (Ref.) | (Ref.) |
|  | 60-69 | 1.21 ([1.19, 1.22], p<0.001) | 1.14 ([1.10, 1.17], p<0.001) | 1.16 ([1.15, 1.18], p<0.001) | 0.98 ([0.97, 1.00], p=0.050) | 1.19 ([1.18, 1.21], p<0.001) |
|  | 70-79 | 1.23 ([1.21, 1.24], p<0.001) | 1.29 ([1.26, 1.32], p<0.001) | 1.12 ([1.10, 1.13], p<0.001) | 0.90 ([0.89, 0.92], p<0.001) | 1.16 ([1.14, 1.17], p<0.001) |
|  | 80-89 | 1.23 ([1.22, 1.25], p<0.001) | 1.29 ([1.25, 1.32], p<0.001) | 1.07 ([1.05, 1.08], p<0.001) | 0.76 ([0.75, 0.78], p<0.001) | 1.02 ([1.01, 1.03], p=0.004) |
|  | 90+ | 1.17 ([1.15, 1.19], p<0.001) | 1.11 ([1.07, 1.15], p<0.001) | 1.03 ([1.01, 1.05], p=0.006) | 0.57 ([0.56, 0.59], p<0.001) | 0.59 ([0.58, 0.60], p<0.001) |
| Sex | Female | (Ref.) | (Ref.) | (Ref.) | (Ref.) | (Ref.) |
|  | Male | 1.01 ([1.00, 1.02], p=0.043) | 1.01 ([0.99, 1.02], p=0.306) | 1.03 ([1.02, 1.03], p<0.001) | 1.07 ([1.06, 1.08], p<0.001) | 1.07 ([1.06, 1.08], p<0.001) |
| IMD 2019 quintiles | 1 (most deprived) | (Ref.) | (Ref.) | (Ref.) | (Ref.) | (Ref.) |
|  | 2 | 0.98 ([0.97, 0.99], p=0.003) | 1.00 ([0.98, 1.02], p=0.903) | 0.98 ([0.97, 1.00], p=0.022) | 1.00 ([0.98, 1.02], p=0.893) | 0.98 ([0.97, 1.00], p=0.011) |
|  | 3 | 0.98 ([0.97, 0.99], p=0.001) | 1.01 ([0.99, 1.03], p=0.435) | 0.98 ([0.96, 0.99], p=0.001) | 1.01 ([0.99, 1.03], p=0.307) | 0.98 ([0.97, 0.99], p=0.001) |
|  | 4 | 0.97 ([0.96, 0.98], p<0.001) | 1.01 ([0.99, 1.03], p=0.509) | 0.97 ([0.96, 0.98], p<0.001) | 1.02 ([1.00, 1.04], p=0.017) | 0.97 ([0.95, 0.98], p<0.001) |
|  | 5 (least deprived) | 0.97 ([0.95, 0.98], p<0.001) | 1.02 ([0.99, 1.04], p=0.156) | 0.96 ([0.95, 0.97], p<0.001) | 1.01 ([0.99, 1.02], p=0.545) | 0.97 ([0.96, 0.98], p<0.001) |
| Ethnicity | White | (Ref.) | (Ref.) | (Ref.) | (Ref.) | (Ref.) |
|  | Asian | 0.93 ([0.91, 0.94], p<0.001) | 0.81 ([0.78, 0.84], p<0.001) | 0.97 ([0.95, 0.98], p<0.001) | 1.08 ([1.05, 1.10], p<0.001) | 0.97 ([0.95, 0.99], p<0.001) |
|  | Black | 0.92 ([0.90, 0.94], p<0.001) | 0.93 ([0.88, 0.98], p=0.003) | 0.94 ([0.92, 0.97], p<0.001) | 1.13 ([1.10, 1.17], p<0.001) | 0.93 ([0.91, 0.95], p<0.001) |
|  | Mixed | 0.88 ([0.85, 0.92], p<0.001) | 0.89 ([0.82, 0.98], p=0.013) | 0.91 ([0.87, 0.96], p<0.001) | 1.00 ([0.95, 1.06], p=0.934) | 0.90 ([0.86, 0.94], p<0.001) |
|  | Other | 0.88 ([0.85, 0.92], p<0.001) | 0.92 ([0.85, 0.99], p=0.031) | 0.91 ([0.87, 0.95], p<0.001) | 0.98 ([0.93, 1.04], p=0.560) | 0.91 ([0.87, 0.95], p<0.001) |
|  | Unknown | 0.88 ([0.82, 0.94], p<0.001) | 1.03 ([0.89, 1.20], p=0.657) | 0.87 ([0.80, 0.94], p=0.001) | 1.10 ([0.99, 1.23], p=0.086) | 0.87 ([0.81, 0.94], p<0.001) |
| Region | East Midlands | (Ref.) | (Ref.) | (Ref.) | (Ref.) | (Ref.) |
|  | East of England | 0.92 ([0.91, 0.94], p<0.001) | 1.05 ([1.02, 1.08], p=0.002) | 0.93 ([0.91, 0.95], p<0.001) | 0.92 ([0.90, 0.95], p<0.001) | 0.94 ([0.92, 0.95], p<0.001) |
|  | London | 0.76 ([0.75, 0.77], p<0.001) | 1.00 ([0.97, 1.04], p=0.822) | 0.78 ([0.77, 0.80], p<0.001) | 0.85 ([0.83, 0.87], p<0.001) | 0.82 ([0.81, 0.84], p<0.001) |
|  | North East | 0.99 ([0.97, 1.01], p=0.564) | 0.92 ([0.89, 0.96], p<0.001) | 1.03 ([1.00, 1.05], p=0.027) | 0.99 ([0.96, 1.02], p=0.570) | 0.98 ([0.96, 1.00], p=0.078) |
|  | North West | 0.98 ([0.96, 0.99], p=0.008) | 0.99 ([0.96, 1.02], p=0.360) | 0.99 ([0.97, 1.00], p=0.136) | 0.97 ([0.95, 0.99], p=0.005) | 0.95 ([0.94, 0.97], p<0.001) |
|  | South East | 0.84 ([0.83, 0.86], p<0.001) | 0.97 ([0.94, 1.00], p=0.032) | 0.88 ([0.87, 0.90], p<0.001) | 0.88 ([0.86, 0.90], p<0.001) | 0.85 ([0.84, 0.87], p<0.001) |
|  | South West | 0.87 ([0.86, 0.89], p<0.001) | 0.93 ([0.91, 0.96], p<0.001) | 0.92 ([0.90, 0.93], p<0.001) | 0.88 ([0.86, 0.90], p<0.001) | 0.85 ([0.83, 0.86], p<0.001) |
|  | West Midlands | 0.88 ([0.86, 0.89], p<0.001) | 0.96 ([0.93, 0.99], p=0.006) | 0.91 ([0.89, 0.92], p<0.001) | 0.90 ([0.88, 0.92], p<0.001) | 0.89 ([0.88, 0.91], p<0.001) |
|  | Yorkshire and The Humber | 0.99 ([0.98, 1.01], p=0.529) | 1.00 ([0.97, 1.04], p=0.776) | 1.00 ([0.98, 1.02], p=0.937) | 0.98 ([0.95, 1.00], p=0.055) | 1.01 ([0.99, 1.02], p=0.425) |
| Charlson comorbidity index | 0 | (Ref.) | (Ref.) | (Ref.) | (Ref.) | (Ref.) |
|  | 1-2 | 1.04 ([1.03, 1.06], p<0.001) | 0.93 ([0.91, 0.96], p<0.001) | 1.12 ([1.10, 1.13], p<0.001) | 1.02 ([1.00, 1.03], p=0.058) | 1.01 ([1.00, 1.03], p=0.017) |
|  | 3-4 | 1.00 ([0.99, 1.02], p=0.453) | 0.91 ([0.89, 0.93], p<0.001) | 1.11 ([1.09, 1.13], p<0.001) | 0.95 ([0.94, 0.97], p<0.001) | 0.95 ([0.94, 0.96], p<0.001) |
|  | 5+ | 0.95 ([0.93, 0.96], p<0.001) | 0.83 ([0.81, 0.85], p<0.001) | 1.11 ([1.09, 1.12], p<0.001) | 0.84 ([0.83, 0.86], p<0.001) | 0.85 ([0.84, 0.86], p<0.001) |
| Data source combination | GDPPR only | (Ref.) | (Ref.) | (Ref.) | (Ref.) | (Ref.) |
|  | HES-APC only | 1.14 ([1.12, 1.16], p<0.001) | 1.35 ([1.31, 1.39], p<0.001) | 0.98 ([0.96, 1.01], p=0.129) | 0.90 ([0.88, 0.93], p<0.001) | 1.06 ([1.04, 1.08], p<0.001) |
|  | SSNAP only | 0.93 ([0.90, 0.96], p<0.001) | 1.25 ([1.18, 1.32], p<0.001) | 0.81 ([0.78, 0.84], p<0.001) | 0.96 ([0.92, 1.00], p=0.066) | 0.94 ([0.91, 0.97], p<0.001) |
|  | GDPPR & HES-APC | 1.67 ([1.64, 1.69], p<0.001) | 1.51 ([1.47, 1.55], p<0.001) | 1.43 ([1.40, 1.45], p<0.001) | 1.07 ([1.05, 1.09], p<0.001) | 1.44 ([1.42, 1.46], p<0.001) |
|  | GDPPR & SSNAP | 1.60 ([1.56, 1.65], p<0.001) | 1.50 ([1.42, 1.58], p<0.001) | 1.39 ([1.34, 1.43], p<0.001) | 1.15 ([1.10, 1.19], p<0.001) | 1.40 ([1.36, 1.44], p<0.001) |
|  | HES-APC & SSNAP | 1.54 ([1.51, 1.56], p<0.001) | 1.53 ([1.49, 1.57], p<0.001) | 1.26 ([1.24, 1.28], p<0.001) | 1.01 ([0.99, 1.03], p=0.328) | 1.37 ([1.35, 1.39], p<0.001) |
|  | GDPPR & HES-APC & SSNAP | 1.74 ([1.72, 1.76], p<0.001) | 1.56 ([1.54, 1.59], p<0.001) | 1.45 ([1.44, 1.47], p<0.001) | 1.15 ([1.13, 1.16], p<0.001) | 1.57 ([1.55, 1.59], p<0.001) |
| Atrial fibrillation | No | (Ref.) | (Ref.) | (Ref.) | (Ref.) | (Ref.) |
|  | Yes | 1.05 ([1.04, 1.06], p<0.001) | 6.74 ([6.61, 6.87], p<0.001) | 0.24 ([0.24, 0.25], p<0.001) | 0.92 ([0.91, 0.94], p<0.001) | 0.93 ([0.92, 0.94], p<0.001) |
| Pre-stroke anticoagulants | No | (Ref.) | (Ref.) | (Ref.) | (Ref.) | (Ref.) |
|  | Yes | 1.19 ([1.18, 1.21], p<0.001) | 3.38 ([3.32, 3.44], p<0.001) | 0.26 ([0.26, 0.27], p<0.001) | 0.95 ([0.93, 0.97], p<0.001) | 0.84 ([0.83, 0.85], p<0.001) |
| Pre-stroke antiplatelets | No | (Ref.) | (Ref.) | (Ref.) | (Ref.) | (Ref.) |
|  | Yes | 1.37 ([1.36, 1.38], p<0.001) | 0.78 ([0.76, 0.79], p<0.001) | 1.64 ([1.62, 1.65], p<0.001) | 0.99 ([0.98, 1.00], p=0.069) | 0.93 ([0.92, 0.93], p<0.001) |
| Pre-stroke antihypertensives | No | (Ref.) | (Ref.) | (Ref.) | (Ref.) | (Ref.) |
|  | Yes | 1.06 ([1.05, 1.06], p<0.001) | 1.04 ([1.03, 1.06], p<0.001) | 1.04 ([1.03, 1.05], p<0.001) | 11.46 ([11.30, 11.61], p<0.001) | 1.03 ([1.02, 1.03], p<0.001) |
| Pre-stroke lipid lowering drugs | No | (Ref.) | (Ref.) | (Ref.) | (Ref.) | (Ref.) |
|  | Yes | 1.05 ([1.04, 1.06], p<0.001) | 1.00 ([0.98, 1.01], p=0.585) | 1.01 ([1.00, 1.02], p=0.022) | 0.99 ([0.97, 1.00], p=0.040) | 2.21 ([2.19, 2.23], p<0.001) |

Table S4.c - Cause-specific multivariable Cox proportional hazard regression for medications dispensed in primary care for non-fatal haemorrhagic strokes (n = 34,435).

|  |  | Hazard ratio ([95% confidence interval], p-value) | | | | |
| --- | --- | --- | --- | --- | --- | --- |
|  |  | Anticoagulants or Antiplatelets | Anticoagulants | Antiplatelets | Antihypertensives | Lipid lowering drugs |
| Year of stroke | 2020 | (Ref.) | (Ref.) | (Ref.) | (Ref.) | (Ref.) |
|  | 2021 | 1.03 ([0.97, 1.10], p=0.317) | 1.05 ([0.97, 1.14], p=0.232) | 1.00 ([0.92, 1.09], p=0.938) | 1.08 ([1.03, 1.13], p=0.001) | 1.08 ([1.03, 1.13], p=0.003) |
|  | 2022 | 1.01 ([0.95, 1.07], p=0.753) | 1.00 ([0.92, 1.09], p=0.937) | 1.05 ([0.97, 1.14], p=0.248) | 1.10 ([1.05, 1.15], p<0.001) | 1.17 ([1.12, 1.23], p<0.001) |
|  | 2023 | 1.06 ([0.99, 1.12], p=0.075) | 1.05 ([0.97, 1.15], p=0.217) | 1.07 ([0.98, 1.17], p=0.113) | 1.08 ([1.04, 1.13], p=0.001) | 1.33 ([1.26, 1.39], p<0.001) |
| Age group (years) | 18-59 | (Ref.) | (Ref.) | (Ref.) | (Ref.) | (Ref.) |
|  | 60-69 | 1.21 ([1.12, 1.30], p<0.001) | 1.14 ([1.02, 1.27], p=0.016) | 1.16 ([1.04, 1.28], p=0.005) | 0.95 ([0.90, 0.99], p=0.024) | 1.20 ([1.14, 1.27], p<0.001) |
|  | 70-79 | 1.13 ([1.05, 1.21], p=0.001) | 0.98 ([0.88, 1.08], p=0.661) | 1.07 ([0.97, 1.18], p=0.151) | 0.85 ([0.81, 0.89], p<0.001) | 1.09 ([1.03, 1.15], p=0.002) |
|  | 80-89 | 1.05 ([0.97, 1.13], p=0.231) | 0.88 ([0.79, 0.98], p=0.020) | 0.99 ([0.89, 1.10], p=0.837) | 0.72 ([0.68, 0.76], p<0.001) | 0.86 ([0.81, 0.91], p<0.001) |
|  | 90+ | 0.83 ([0.75, 0.93], p=0.001) | 0.62 ([0.53, 0.72], p<0.001) | 0.85 ([0.73, 0.99], p=0.038) | 0.53 ([0.49, 0.58], p<0.001) | 0.50 ([0.45, 0.55], p<0.001) |
| Sex | Female | (Ref.) | (Ref.) | (Ref.) | (Ref.) | (Ref.) |
|  | Male | 1.04 ([0.99, 1.08], p=0.124) | 0.98 ([0.93, 1.05], p=0.602) | 1.12 ([1.05, 1.19], p<0.001) | 1.16 ([1.13, 1.20], p<0.001) | 1.06 ([1.02, 1.10], p=0.001) |
| IMD 2019 quintiles | 1 (most deprived) | (Ref.) | (Ref.) | (Ref.) | (Ref.) | (Ref.) |
|  | 2 | 0.97 ([0.91, 1.04], p=0.425) | 0.96 ([0.87, 1.06], p=0.373) | 0.97 ([0.89, 1.07], p=0.591) | 1.03 ([0.98, 1.08], p=0.290) | 1.00 ([0.94, 1.05], p=0.889) |
|  | 3 | 1.01 ([0.94, 1.08], p=0.808) | 1.00 ([0.91, 1.10], p=0.987) | 1.01 ([0.91, 1.11], p=0.917) | 1.05 ([0.99, 1.10], p=0.090) | 1.02 ([0.97, 1.08], p=0.383) |
|  | 4 | 0.97 ([0.90, 1.04], p=0.341) | 0.97 ([0.88, 1.07], p=0.572) | 0.96 ([0.87, 1.05], p=0.370) | 1.03 ([0.98, 1.09], p=0.214) | 1.01 ([0.96, 1.07], p=0.600) |
|  | 5 (least deprived) | 0.95 ([0.88, 1.02], p=0.135) | 0.96 ([0.87, 1.06], p=0.463) | 0.92 ([0.83, 1.02], p=0.107) | 1.03 ([0.98, 1.08], p=0.290) | 1.00 ([0.94, 1.05], p=0.926) |
| Ethnicity | White | (Ref.) | (Ref.) | (Ref.) | (Ref.) | (Ref.) |
|  | Asian | 0.98 ([0.90, 1.07], p=0.668) | 0.87 ([0.76, 1.01], p=0.066) | 1.08 ([0.97, 1.21], p=0.172) | 1.19 ([1.12, 1.26], p<0.001) | 1.15 ([1.08, 1.22], p<0.001) |
|  | Black | 1.00 ([0.88, 1.14], p=0.955) | 0.86 ([0.70, 1.06], p=0.152) | 1.13 ([0.96, 1.33], p=0.134) | 1.33 ([1.23, 1.43], p<0.001) | 1.11 ([1.01, 1.22], p=0.032) |
|  | Mixed | 0.87 ([0.68, 1.10], p=0.245) | 0.59 ([0.38, 0.90], p=0.016) | 1.09 ([0.82, 1.45], p=0.544) | 1.08 ([0.93, 1.24], p=0.306) | 0.85 ([0.71, 1.01], p=0.069) |
|  | Other | 0.93 ([0.76, 1.15], p=0.500) | 0.82 ([0.60, 1.14], p=0.237) | 1.03 ([0.79, 1.33], p=0.825) | 1.18 ([1.04, 1.35], p=0.011) | 1.15 ([1.00, 1.33], p=0.057) |
|  | Unknown | 0.80 ([0.45, 1.41], p=0.441) | 1.10 ([0.55, 2.21], p=0.784) | 0.64 ([0.27, 1.55], p=0.325) | 1.66 ([1.27, 2.17], p<0.001) | 1.05 ([0.70, 1.56], p=0.829) |
| Region | East Midlands | (Ref.) | (Ref.) | (Ref.) | (Ref.) | (Ref.) |
|  | East of England | 0.94 ([0.85, 1.03], p=0.173) | 0.95 ([0.84, 1.08], p=0.438) | 0.96 ([0.84, 1.10], p=0.588) | 0.99 ([0.93, 1.07], p=0.874) | 0.92 ([0.86, 0.99], p=0.034) |
|  | London | 0.95 ([0.86, 1.05], p=0.311) | 0.88 ([0.77, 1.00], p=0.059) | 1.09 ([0.95, 1.24], p=0.214) | 0.89 ([0.83, 0.95], p=0.001) | 0.94 ([0.88, 1.02], p=0.118) |
|  | North East | 0.88 ([0.78, 1.00], p=0.043) | 0.80 ([0.68, 0.94], p=0.006) | 0.89 ([0.76, 1.06], p=0.197) | 1.08 ([0.99, 1.17], p=0.084) | 1.12 ([1.03, 1.23], p=0.011) |
|  | North West | 1.01 ([0.93, 1.11], p=0.762) | 0.95 ([0.84, 1.07], p=0.413) | 1.07 ([0.94, 1.21], p=0.283) | 0.96 ([0.90, 1.03], p=0.249) | 0.96 ([0.90, 1.03], p=0.298) |
|  | South East | 1.00 ([0.91, 1.09], p=0.978) | 0.96 ([0.86, 1.09], p=0.549) | 1.03 ([0.91, 1.16], p=0.668) | 0.88 ([0.83, 0.94], p<0.001) | 0.99 ([0.92, 1.06], p=0.741) |
|  | South West | 1.01 ([0.92, 1.11], p=0.878) | 0.98 ([0.86, 1.11], p=0.740) | 1.01 ([0.88, 1.16], p=0.866) | 0.89 ([0.83, 0.96], p=0.002) | 0.97 ([0.90, 1.04], p=0.361) |
|  | West Midlands | 0.97 ([0.88, 1.07], p=0.528) | 0.93 ([0.81, 1.06], p=0.281) | 0.99 ([0.87, 1.14], p=0.931) | 0.95 ([0.88, 1.02], p=0.151) | 0.97 ([0.90, 1.04], p=0.372) |
|  | Yorkshire and The Humber | 1.00 ([0.90, 1.10], p=0.922) | 0.87 ([0.76, 1.00], p=0.054) | 1.08 ([0.94, 1.24], p=0.270) | 1.07 ([1.00, 1.15], p=0.059) | 1.17 ([1.08, 1.26], p<0.001) |
| Charlson comorbidity index | 0 | (Ref.) | (Ref.) | (Ref.) | (Ref.) | (Ref.) |
|  | 1-2 | 1.09 ([1.01, 1.17], p=0.024) | 1.00 ([0.90, 1.11], p=0.970) | 1.14 ([1.03, 1.26], p=0.014) | 1.01 ([0.96, 1.06], p=0.792) | 1.07 ([1.01, 1.13], p=0.015) |
|  | 3-4 | 1.13 ([1.05, 1.23], p=0.002) | 1.05 ([0.94, 1.17], p=0.390) | 1.20 ([1.08, 1.34], p=0.001) | 1.00 ([0.94, 1.05], p=0.871) | 0.99 ([0.94, 1.05], p=0.764) |
|  | 5+ | 1.15 ([1.06, 1.25], p=0.001) | 1.04 ([0.93, 1.17], p=0.466) | 1.19 ([1.06, 1.33], p=0.003) | 0.85 ([0.80, 0.90], p<0.001) | 0.91 ([0.86, 0.97], p=0.005) |
| Data source combination | GDPPR only | (Ref.) | (Ref.) | (Ref.) | (Ref.) | (Ref.) |
|  | HES-APC only | 0.79 ([0.73, 0.85], p<0.001) | 0.94 ([0.84, 1.04], p=0.227) | 0.67 ([0.60, 0.75], p<0.001) | 1.34 ([1.25, 1.44], p<0.001) | 0.88 ([0.83, 0.95], p<0.001) |
|  | SSNAP only | 0.72 ([0.63, 0.83], p<0.001) | 0.90 ([0.75, 1.08], p=0.254) | 0.60 ([0.49, 0.73], p<0.001) | 1.53 ([1.37, 1.71], p<0.001) | 0.94 ([0.84, 1.06], p=0.313) |
|  | GDPPR & HES-APC | 0.71 ([0.66, 0.77], p<0.001) | 0.81 ([0.73, 0.90], p<0.001) | 0.67 ([0.60, 0.74], p<0.001) | 1.84 ([1.74, 1.96], p<0.001) | 1.06 ([1.00, 1.13], p=0.042) |
|  | GDPPR & SSNAP | 0.71 ([0.62, 0.81], p<0.001) | 0.78 ([0.65, 0.94], p=0.007) | 0.67 ([0.55, 0.81], p<0.001) | 2.12 ([1.92, 2.33], p<0.001) | 1.19 ([1.08, 1.32], p=0.001) |
|  | HES-APC & SSNAP | 0.65 ([0.60, 0.70], p<0.001) | 0.72 ([0.65, 0.79], p<0.001) | 0.62 ([0.56, 0.69], p<0.001) | 2.03 ([1.91, 2.15], p<0.001) | 1.04 ([0.98, 1.10], p=0.204) |
|  | GDPPR & HES-APC & SSNAP | 0.67 ([0.63, 0.71], p<0.001) | 0.70 ([0.64, 0.75], p<0.001) | 0.66 ([0.61, 0.72], p<0.001) | 2.32 ([2.21, 2.43], p<0.001) | 1.20 ([1.15, 1.26], p<0.001) |
| Atrial fibrillation | No | (Ref.) | (Ref.) | (Ref.) | (Ref.) | (Ref.) |
|  | Yes | 1.28 ([1.20, 1.37], p<0.001) | 1.57 ([1.43, 1.73], p<0.001) | 0.94 ([0.85, 1.04], p=0.215) | 1.01 ([0.95, 1.07], p=0.730) | 0.98 ([0.93, 1.04], p=0.525) |
| Pre-stroke anticoagulants | No | (Ref.) | (Ref.) | (Ref.) | (Ref.) | (Ref.) |
|  | Yes | 3.42 ([3.20, 3.66], p<0.001) | 8.79 ([8.00, 9.67], p<0.001) | 0.75 ([0.67, 0.85], p<0.001) | 0.89 ([0.84, 0.94], p<0.001) | 0.98 ([0.93, 1.05], p=0.627) |
| Pre-stroke antiplatelets | No | (Ref.) | (Ref.) | (Ref.) | (Ref.) | (Ref.) |
|  | Yes | 3.25 ([3.09, 3.42], p<0.001) | 1.07 ([0.98, 1.17], p=0.131) | 7.25 ([6.73, 7.81], p<0.001) | 0.97 ([0.93, 1.01], p=0.179) | 1.09 ([1.04, 1.13], p<0.001) |
| Pre-stroke antihypertensives | No | (Ref.) | (Ref.) | (Ref.) | (Ref.) | (Ref.) |
|  | Yes | 1.16 ([1.10, 1.21], p<0.001) | 0.98 ([0.92, 1.04], p=0.435) | 1.28 ([1.20, 1.37], p<0.001) | 5.04 ([4.86, 5.22], p<0.001) | 1.09 ([1.05, 1.13], p<0.001) |
| Pre-stroke lipid lowering drugs | No | (Ref.) | (Ref.) | (Ref.) | (Ref.) | (Ref.) |
|  | Yes | 1.17 ([1.12, 1.24], p<0.001) | 1.01 ([0.94, 1.08], p=0.767) | 1.23 ([1.14, 1.33], p<0.001) | 1.02 ([0.98, 1.05], p=0.431) | 8.70 ([8.31, 9.09], p<0.001) |

Table S5.a - Unadjusted and covariate adjusted mean home-time in the 180-days following stroke onset in non-fatal strokes and hospital survivors identified in GDPPR, HES-APC and SSNAP.

|  |  | **All stroke types (n=353,325)** | | **Ischaemic or unknown stroke type**  **(n=318,160)** | | **Haemorrhagic strokes (n=35,165)** | |
| --- | --- | --- | --- | --- | --- | --- | --- |
|  |  | Unadjusted | Adjusted | Unadjusted | Adjusted | Unadjusted | Adjusted |
| Overall | Overall | 166.6 (166.4, 166.7) | - | 167.1 (167.0, 167.2) | - | 161.6 (161.2, 162.1) | - |
| Year of stroke | 2020 | 164.4 (164.1, 164.7) | 163.9 (163.7, 164.1) | 165.0 (164.8, 165.3) | 164.6 (164.3, 164.8) | 158.6 (157.6, 159.6) | 158.0 (157.1, 158.9) |
|  | 2021 | 165.5 (165.3, 165.8) | 165.4 (165.2, 165.6) | 166.2 (165.9, 166.4) | 166.1 (165.8, 166.3) | 159.5 (158.6, 160.5) | 159.6 (158.7, 160.4) |
|  | 2022 | 166.8 (166.5, 167.0) | 166.9 (166.6, 167.1) | 167.2 (166.9, 167.5) | 167.3 (167.0, 167.5) | 162.9 (162.0, 163.8) | 163.0 (162.1, 163.8) |
|  | 2023 | 169.3 (169.0, 169.5) | 169.7 (169.5, 169.9) | 169.8 (169.5, 170.0) | 170.2 (170.0, 170.4) | 164.9 (164.1, 165.8) | 165.3 (164.4, 166.1) |
| Age group (years) | 18-59 | 176.8 (176.7, 176.9) | 173.9 (173.7, 174.2) | 177.2 (177.1, 177.3) | 174.1 (173.9, 174.4) | 174.1 (173.6, 174.6) | 172.2 (171.3, 173.0) |
|  | 60-69 | 173.6 (173.4, 173.8) | 171.9 (171.6, 172.1) | 174.1 (173.9, 174.3) | 172.4 (172.1, 172.6) | 168.3 (167.4, 169.2) | 167.2 (166.2, 168.2) |
|  | 70-79 | 168.5 (168.3, 168.7) | 168.3 (168.1, 168.6) | 169.3 (169.0, 169.5) | 169.0 (168.8, 169.3) | 161.1 (160.2, 162.1) | 161.2 (160.3, 162.0) |
|  | 80-89 | 157.6 (157.3, 157.9) | 160.1 (159.9, 160.4) | 158.5 (158.2, 158.8) | 161.0 (160.8, 161.3) | 149.1 (148.0, 150.3) | 151.3 (150.4, 152.2) |
|  | 90+ | 141.4 (140.7, 142.1) | 146.4 (145.9, 146.8) | 141.9 (141.2, 142.6) | 147.1 (146.7, 147.6) | 135.9 (133.4, 138.4) | 139.3 (137.6, 141.1) |
| Sex | Female | 164.7 (164.5, 164.9) | 166.9 (166.7, 167.1) | 165.2 (165.0, 165.4) | 167.4 (167.3, 167.6) | 159.9 (159.2, 160.6) | 162.2 (161.6, 162.9) |
|  | Male | 168.2 (168.0, 168.3) | 166.3 (166.1, 166.4) | 168.7 (168.6, 168.9) | 166.9 (166.7, 167.0) | 163.1 (162.5, 163.7) | 161.1 (160.5, 161.7) |
| IMD 2019 quintiles | 1 (most deprived) | 167.0 (166.7, 167.2) | 165.7 (165.4, 166.0) | 167.5 (167.2, 167.8) | 166.3 (166.1, 166.6) | 162.0 (160.9, 163.0) | 159.9 (158.9, 160.9) |
|  | 2 | 166.6 (166.3, 166.9) | 166.1 (165.9, 166.4) | 167.1 (166.8, 167.4) | 166.7 (166.4, 167.0) | 161.9 (160.9, 162.9) | 161.0 (160.0, 162.0) |
|  | 3 | 166.5 (166.2, 166.8) | 166.7 (166.4, 166.9) | 167.1 (166.8, 167.3) | 167.2 (167.0, 167.5) | 161.6 (160.6, 162.6) | 161.7 (160.8, 162.7) |
|  | 4 | 166.4 (166.1, 166.6) | 167.0 (166.8, 167.3) | 167.0 (166.7, 167.2) | 167.6 (167.3, 167.8) | 161.2 (160.2, 162.2) | 162.2 (161.2, 163.1) |
|  | 5 (least deprived) | 166.4 (166.1, 166.7) | 167.3 (167.0, 167.6) | 167.0 (166.7, 167.3) | 167.8 (167.5, 168.1) | 161.4 (160.4, 162.5) | 163.2 (162.2, 164.2) |
| Ethnicity | White | 166.0 (165.9, 166.2) | 166.3 (166.2, 166.4) | 166.6 (166.5, 166.8) | 166.9 (166.8, 167.0) | 160.4 (159.9, 160.9) | 161.0 (160.5, 161.5) |
|  | Asian | 171.6 (171.1, 172.0) | 169.3 (168.8, 169.8) | 172.0 (171.6, 172.4) | 169.6 (169.1, 170.1) | 168.7 (167.2, 170.1) | 165.9 (164.2, 167.5) |
|  | Black | 170.8 (170.2, 171.4) | 169.1 (168.4, 169.8) | 170.8 (170.2, 171.5) | 169.3 (168.5, 170.0) | 170.4 (168.7, 172.2) | 165.9 (163.7, 168.1) |
|  | Mixed | 170.3 (169.1, 171.5) | 167.4 (166.1, 168.6) | 170.5 (169.2, 171.7) | 167.7 (166.3, 169.0) | 169.4 (165.7, 173.0) | 163.7 (159.6, 167.7) |
|  | Other | 170.4 (169.3, 171.4) | 167.5 (166.4, 168.6) | 170.8 (169.7, 171.9) | 167.8 (166.7, 169.0) | 167.2 (163.8, 170.7) | 163.9 (160.2, 167.6) |
|  | Unknown | 166.4 (163.9, 168.8) | 161.9 (159.8, 164.0) | 166.6 (164.0, 169.1) | 162.3 (160.2, 164.5) | 164.4 (155.9, 172.9) | 157.4 (149.4, 165.5) |
| Region | East Midlands | 167.0 (166.6, 167.4) | 166.5 (166.1, 166.9) | 167.5 (167.1, 167.9) | 166.9 (166.5, 167.3) | 162.6 (161.2, 164.1) | 162.6 (161.1, 164.0) |
|  | East of England | 165.6 (165.2, 166.0) | 165.6 (165.3, 166.0) | 166.2 (165.8, 166.6) | 166.2 (165.8, 166.5) | 160.5 (159.1, 161.9) | 160.8 (159.5, 162.0) |
|  | London | 167.6 (167.3, 168.0) | 167.5 (167.1, 167.8) | 168.2 (167.8, 168.5) | 168.1 (167.7, 168.5) | 163.3 (162.1, 164.6) | 161.6 (160.4, 162.9) |
|  | North East | 167.0 (166.5, 167.5) | 166.9 (166.4, 167.4) | 167.6 (167.1, 168.1) | 167.3 (166.8, 167.8) | 161.9 (159.9, 163.8) | 163.1 (161.2, 165.0) |
|  | North West | 166.1 (165.8, 166.4) | 166.6 (166.3, 166.9) | 166.6 (166.3, 167.0) | 167.2 (166.9, 167.5) | 161.4 (160.2, 162.6) | 161.5 (160.4, 162.7) |
|  | South East | 166.9 (166.6, 167.2) | 166.9 (166.6, 167.2) | 167.5 (167.2, 167.8) | 167.5 (167.2, 167.8) | 161.4 (160.2, 162.5) | 161.8 (160.8, 162.9) |
|  | South West | 166.3 (165.9, 166.6) | 166.4 (166.1, 166.7) | 166.8 (166.4, 167.1) | 166.9 (166.6, 167.3) | 161.6 (160.2, 163.0) | 161.8 (160.5, 163.1) |
|  | West Midlands | 165.9 (165.4, 166.3) | 166.2 (165.8, 166.5) | 166.4 (166.0, 166.8) | 166.8 (166.4, 167.2) | 160.7 (159.2, 162.1) | 160.9 (159.6, 162.2) |
|  | Yorkshire and The Humber | 166.9 (166.5, 167.2) | 166.4 (166.0, 166.7) | 167.4 (167.0, 167.8) | 166.9 (166.5, 167.2) | 161.2 (159.6, 162.7) | 161.1 (159.7, 162.5) |
| Charlson comorbidity index | 0 | 175.5 (175.4, 175.7) | 170.1 (169.8, 170.4) | 175.9 (175.7, 176.0) | 170.4 (170.1, 170.7) | 172.1 (171.3, 172.9) | 166.4 (165.2, 167.5) |
|  | 1-2 | 171.5 (171.4, 171.7) | 170.1 (169.9, 170.2) | 172.0 (171.9, 172.2) | 170.5 (170.3, 170.7) | 167.5 (166.9, 168.1) | 166.0 (165.3, 166.6) |
|  | 3-4 | 164.3 (164.0, 164.6) | 166.9 (166.6, 167.1) | 164.9 (164.6, 165.2) | 167.4 (167.2, 167.7) | 159.3 (158.4, 160.2) | 161.7 (160.8, 162.5) |
|  | 5+ | 150.2 (149.8, 150.6) | 155.4 (155.1, 155.6) | 151.2 (150.8, 151.6) | 156.3 (156.1, 156.6) | 141.0 (139.5, 142.4) | 146.5 (145.4, 147.5) |
| Data source combination | GDPPR only | 173.8 (173.6, 173.9) | 171.9 (168.6, 175.2) | 174.2 (174.1, 174.4) | 172.1 (168.7, 175.5) | 167.6 (166.8, 168.4) | 168.9 (157.1, 180.8) |
|  | HES-APC only | 151.5 (150.8, 152.2) | 153.9 (150.5, 157.2) | 151.8 (151.0, 152.6) | 154.3 (150.8, 157.7) | 150.2 (148.5, 151.9) | 154.2 (142.3, 166.1) |
|  | SSNAP only | 163.8 (162.8, 164.8) | 162.2 (159.2, 165.1) | 165.3 (164.2, 166.3) | 163.2 (160.2, 166.3) | 155.6 (152.5, 158.6) | 154.8 (144.2, 165.4) |
|  | GDPPR & HES-APC | 163.4 (162.9, 163.8) | 164.6 (161.3, 167.9) | 163.9 (163.4, 164.3) | 165.0 (161.5, 168.4) | 159.8 (158.4, 161.1) | 162.4 (150.5, 174.3) |
|  | GDPPR & SSNAP | 169.5 (168.7, 170.2) | 166.5 (163.5, 169.4) | 170.5 (169.7, 171.3) | 167.2 (164.2, 170.3) | 163.6 (161.1, 166.0) | 160.5 (149.9, 171.0) |
|  | HES-APC & SSNAP | 154.3 (153.8, 154.9) | 158.5 (155.7, 161.4) | 154.3 (153.7, 154.9) | 158.8 (155.9, 161.7) | 154.6 (153.1, 156.1) | 155.3 (145.0, 165.6) |
|  | GDPPR & HES-APC & SSNAP | 167.6 (167.4, 167.8) | 167.4 (164.6, 170.2) | 167.8 (167.6, 168.0) | 167.9 (165.0, 170.8) | 165.4 (164.7, 166.0) | 162.3 (152.0, 172.5) |
| NIHSS score on arrival | 0-4 | 171.0 (170.8, 171.2) | 168.4 (165.5, 171.2) | 171.2 (171.0, 171.4) | 168.5 (165.6, 171.4) | 168.5 (167.8, 169.1) | 167.4 (157.2, 177.7) |
|  | 5-10 | 162.7 (162.3, 163.0) | 165.1 (162.2, 167.9) | 162.9 (162.5, 163.3) | 165.3 (162.4, 168.2) | 160.3 (159.0, 161.5) | 163.4 (153.1, 173.7) |
|  | 11-15 | 152.3 (151.5, 153.2) | 158.5 (155.6, 161.4) | 151.8 (150.8, 152.7) | 158.4 (155.4, 161.3) | 155.3 (153.2, 157.3) | 159.2 (148.8, 169.5) |
|  | 16-21 | 145.9 (144.8, 147.0) | 153.6 (150.8, 156.5) | 144.8 (143.6, 146.0) | 153.1 (150.1, 156.1) | 151.3 (148.8, 153.8) | 155.6 (145.1, 166.0) |
|  | 22+ | 137.4 (135.8, 139.1) | 146.7 (143.7, 149.7) | 135.1 (133.3, 137.0) | 145.2 (142.1, 148.3) | 149.5 (145.9, 153.2) | 154.1 (143.5, 164.6) |
|  | (missing) | 168.4 (168.2, 168.6) | 168.1 (164.8, 171.4) | 169.2 (169.1, 169.4) | 169.0 (165.6, 172.4) | 161.0 (160.3, 161.7) | 159.3 (147.5, 171.2) |
| Rankin score at discharge | 0 | 176.6 (176.3, 176.8) | 173.5 (172.9, 174.0) | 176.7 (176.4, 177.0) | 174.0 (173.5, 174.5) | 174.6 (173.2, 176.0) | 167.1 (164.6, 169.5) |
|  | 1 | 176.3 (176.1, 176.5) | 174.3 (173.9, 174.7) | 176.3 (176.1, 176.5) | 174.8 (174.3, 175.2) | 176.3 (175.5, 177.1) | 169.3 (167.4, 171.3) |
|  | 2 | 174.0 (173.7, 174.3) | 174.1 (173.7, 174.6) | 174.0 (173.7, 174.3) | 174.5 (174.1, 175.0) | 174.3 (173.3, 175.3) | 170.1 (168.1, 172.0) |
|  | 3 | 166.4 (165.9, 166.9) | 171.7 (171.2, 172.2) | 166.4 (165.9, 166.9) | 172.2 (171.6, 172.7) | 165.9 (164.4, 167.5) | 166.8 (165.0, 168.7) |
|  | 4 | 151.8 (151.0, 152.5) | 161.7 (161.2, 162.3) | 151.3 (150.4, 152.1) | 161.8 (161.3, 162.4) | 155.4 (153.4, 157.4) | 160.3 (158.5, 162.1) |
|  | 5 | 126.7 (125.0, 128.3) | 142.1 (141.2, 142.9) | 124.7 (122.9, 126.5) | 141.4 (140.5, 142.3) | 136.4 (132.6, 140.2) | 144.2 (141.7, 146.8) |
|  | (missing) | 165.8 (165.6, 165.9) | 164.7 (164.6, 164.9) | 166.5 (166.3, 166.6) | 165.2 (165.0, 165.4) | 160.1 (159.6, 160.7) | 160.5 (160.0, 161.1) |

Table S5.b - Multivariable linear regression model of home-time in the 180-days following stroke onset in non-fatal strokes and hospital survivors identified in GDPPR, HES-APC and SSNAP.

|  |  | **All stroke types (n = 353,325)** | **Ischaemic or unknown stroke type**  **(n=318,160)** | **Haemorrhagic strokes**  **(n=35,165)** |
| --- | --- | --- | --- | --- |
| (Intercept) | (Intercept) | 187.89 ([181.7, 194.07], p<0.001) | 187.26 ([180.87, 193.65], p<0.001) | 191.18 ([168.81, 213.54], p<0.001) |
| Year of stroke | 2020 | (Ref.) | (Ref.) | (Ref.) |
|  | 2021 | 1.51 ([1.18, 1.84], p<0.001) | 1.49 ([1.15, 1.83], p<0.001) | 1.51 ([0.28, 2.74], p=0.017) |
|  | 2022 | 2.95 ([2.62, 3.28], p<0.001) | 2.71 ([2.37, 3.05], p<0.001) | 4.94 ([3.71, 6.16], p<0.001) |
|  | 2023 | 5.81 ([5.48, 6.13], p<0.001) | 5.63 ([5.3, 5.97], p<0.001) | 7.22 ([6.01, 8.44], p<0.001) |
| Age group (years) | 18-59 | (Ref.) | (Ref.) | (Ref.) |
|  | 60-69 | -2.05 ([-2.42, -1.69], p<0.001) | -1.78 ([-2.15, -1.4], p<0.001) | -4.99 ([-6.31, -3.67], p<0.001) |
|  | 70-79 | -5.58 ([-5.92, -5.24], p<0.001) | -5.08 ([-5.43, -4.73], p<0.001) | -11.02 ([-12.24, -9.79], p<0.001) |
|  | 80-89 | -13.8 ([-14.16, -13.44], p<0.001) | -13.09 ([-13.46, -12.72], p<0.001) | -20.82 ([-22.12, -19.53], p<0.001) |
|  | 90+ | -27.56 ([-28.08, -27.05], p<0.001) | -27.02 ([-27.55, -26.49], p<0.001) | -32.82 ([-34.82, -30.83], p<0.001) |
| Sex | Female | (Ref.) | (Ref.) | (Ref.) |
|  | Male | -0.62 ([-0.85, -0.39], p<0.001) | -0.57 ([-0.81, -0.33], p<0.001) | -1.15 ([-2.02, -0.28], p=0.009) |
| IMD 2019 quintiles | 1 (most deprived) | (Ref.) | (Ref.) | (Ref.) |
|  | 2 | 0.42 ([0.06, 0.79], p=0.023) | 0.35 ([-0.03, 0.72], p=0.072) | 1.11 ([-0.27, 2.5], p=0.115) |
|  | 3 | 0.97 ([0.6, 1.34], p<0.001) | 0.88 ([0.5, 1.26], p<0.001) | 1.81 ([0.41, 3.2], p=0.011) |
|  | 4 | 1.33 ([0.95, 1.7], p<0.001) | 1.24 ([0.86, 1.62], p<0.001) | 2.27 ([0.87, 3.66], p=0.001) |
|  | 5 (least deprived) | 1.6 ([1.22, 1.99], p<0.001) | 1.44 ([1.05, 1.84], p<0.001) | 3.3 ([1.87, 4.72], p<0.001) |
| Ethnicity | White | (Ref.) | (Ref.) | (Ref.) |
|  | Asian | 3.03 ([2.52, 3.54], p<0.001) | 2.71 ([2.18, 3.24], p<0.001) | 4.86 ([3.15, 6.58], p<0.001) |
|  | Black | 2.81 ([2.12, 3.51], p<0.001) | 2.36 ([1.64, 3.09], p<0.001) | 4.9 ([2.62, 7.18], p<0.001) |
|  | Mixed | 1.07 ([-0.19, 2.34], p=0.096) | 0.77 ([-0.56, 2.09], p=0.257) | 2.69 ([-1.4, 6.78], p=0.197) |
|  | Other | 1.21 ([0.1, 2.33], p=0.033) | 0.91 ([-0.25, 2.08], p=0.124) | 2.92 ([-0.78, 6.62], p=0.122) |
|  | Unknown | -4.4 ([-6.5, -2.3], p<0.001) | -4.58 ([-6.74, -2.42], p<0.001) | -3.55 ([-11.6, 4.51], p=0.388) |
| Region | East Midlands | (Ref.) | (Ref.) | (Ref.) |
|  | East of England | -0.88 ([-1.39, -0.37], p=0.001) | -0.73 ([-1.26, -0.21], p=0.006) | -1.81 ([-3.72, 0.1], p=0.063) |
|  | London | 0.96 ([0.44, 1.49], p<0.001) | 1.19 ([0.65, 1.73], p<0.001) | -0.95 ([-2.84, 0.95], p=0.329) |
|  | North East | 0.39 ([-0.22, 1], p=0.21) | 0.4 ([-0.23, 1.02], p=0.214) | 0.54 ([-1.8, 2.88], p=0.65) |
|  | North West | 0.14 ([-0.34, 0.63], p=0.567) | 0.3 ([-0.2, 0.8], p=0.237) | -1.02 ([-2.84, 0.8], p=0.274) |
|  | South East | 0.43 ([-0.04, 0.91], p=0.071) | 0.61 ([0.13, 1.1], p=0.014) | -0.72 ([-2.49, 1.04], p=0.421) |
|  | South West | -0.09 ([-0.59, 0.42], p=0.735) | 0 ([-0.52, 0.52], p=0.998) | -0.77 ([-2.69, 1.14], p=0.428) |
|  | West Midlands | -0.32 ([-0.84, 0.21], p=0.234) | -0.13 ([-0.67, 0.41], p=0.635) | -1.66 ([-3.62, 0.3], p=0.097) |
|  | Yorkshire and The Humber | -0.14 ([-0.65, 0.37], p=0.596) | -0.05 ([-0.57, 0.48], p=0.862) | -1.42 ([-3.42, 0.58], p=0.163) |
| Charlson comorbidity index | 0 | (Ref.) | (Ref.) | (Ref.) |
|  | 1-2 | -0.02 ([-0.35, 0.32], p=0.928) | 0.1 ([-0.25, 0.45], p=0.581) | -0.4 ([-1.69, 0.9], p=0.547) |
|  | 3-4 | -3.22 ([-3.6, -2.85], p<0.001) | -3 ([-3.38, -2.61], p<0.001) | -4.69 ([-6.11, -3.27], p<0.001) |
|  | 5+ | -14.71 ([-15.12, -14.31], p<0.001) | -14.08 ([-14.5, -13.66], p<0.001) | -19.9 ([-21.46, -18.33], p<0.001) |
| Data source combination | GDPPR only | (Ref.) | (Ref.) | (Ref.) |
|  | HES-APC only | -18.02 ([-18.54, -17.49], p<0.001) | -17.82 ([-18.38, -17.26], p<0.001) | -14.75 ([-16.35, -13.16], p<0.001) |
|  | SSNAP only | -9.74 ([-15.93, -3.55], p=0.002) | -8.88 ([-15.27, -2.48], p=0.007) | -14.13 ([-36.37, 8.11], p=0.213) |
|  | GDPPR & HES-APC | -7.3 ([-7.72, -6.88], p<0.001) | -7.09 ([-7.53, -6.66], p<0.001) | -6.55 ([-8.05, -5.05], p<0.001) |
|  | GDPPR & SSNAP | -5.44 ([-11.63, 0.75], p=0.085) | -4.85 ([-11.25, 1.55], p=0.138) | -8.45 ([-30.68, 13.78], p=0.456) |
|  | HES-APC & SSNAP | -13.37 ([-19.52, -7.23], p<0.001) | -13.27 ([-19.62, -6.92], p<0.001) | -13.65 ([-35.77, 8.47], p=0.227) |
|  | GDPPR & HES-APC & SSNAP | -4.49 ([-10.63, 1.65], p=0.152) | -4.16 ([-10.5, 2.19], p=0.199) | -6.66 ([-28.78, 15.46], p=0.555) |
| NIHSS score on arrival | 0-4 | (Ref.) | (Ref.) | (Ref.) |
|  | 5-10 | -3.29 ([-3.67, -2.91], p<0.001) | -3.23 ([-3.61, -2.84], p<0.001) | -4.02 ([-5.48, -2.55], p<0.001) |
|  | 11-15 | -9.86 ([-10.47, -9.24], p<0.001) | -10.18 ([-10.83, -9.52], p<0.001) | -8.28 ([-10.23, -6.33], p<0.001) |
|  | 16-21 | -14.72 ([-15.42, -14.02], p<0.001) | -15.4 ([-16.14, -14.65], p<0.001) | -11.88 ([-14.04, -9.72], p<0.001) |
|  | 22+ | -21.66 ([-22.6, -20.72], p<0.001) | -23.33 ([-24.33, -22.33], p<0.001) | -13.38 ([-16.23, -10.53], p<0.001) |
|  | (missing) | -0.29 ([-6.42, 5.84], p=0.925) | 0.47 ([-5.87, 6.8], p=0.885) | -8.1 ([-30.17, 13.98], p=0.472) |
| Rankin score at discharge | 0 | (Ref.) | (Ref.) | (Ref.) |
|  | 1 | 0.86 ([0.24, 1.47], p=0.006) | 0.74 ([0.12, 1.36], p=0.019) | 2.26 ([-0.69, 5.2], p=0.133) |
|  | 2 | 0.67 ([0.02, 1.31], p=0.044) | 0.5 ([-0.16, 1.15], p=0.136) | 2.98 ([0, 5.96], p=0.05) |
|  | 3 | -1.8 ([-2.48, -1.11], p<0.001) | -1.86 ([-2.56, -1.16], p<0.001) | -0.24 ([-3.23, 2.75], p=0.873) |
|  | 4 | -11.73 ([-12.47, -11], p<0.001) | -12.18 ([-12.94, -11.42], p<0.001) | -6.81 ([-9.81, -3.81], p<0.001) |
|  | 5 | -31.39 ([-32.4, -30.38], p<0.001) | -32.61 ([-33.68, -31.55], p<0.001) | -22.85 ([-26.38, -19.32], p<0.001) |
|  | (missing) | -8.74 ([-9.3, -8.18], p<0.001) | -8.84 ([-9.41, -8.27], p<0.001) | -6.55 ([-9.13, -3.97], p<0.001) |

Table S6.a - Codelists and data sources used to define study variables and phenotypes

| **Variable** | **Data sources** | **Codelist source or code matching logic** | **Additional specifications** |
| --- | --- | --- | --- |
| Stroke | GDPPR, HES-APC, SSNAP | <https://phenotypes.healthdatagateway.org/phenotypes/PH983/version/2161/detail/> |  |
| Smoking status | GDPPR | <https://www.opencodelists.org/codelist/nhsd-primary-care-domain-refsets/exsmok_cod/20200812/> |  |
| Body mass index | GDPPR | <https://www.opencodelists.org/codelist/nhsd-primary-care-domain-refsets/bmival_cod/20201016/> | Minimum: 5; Maximum: 100 |
| Systolic blood pressure | GDPPR | <https://www.opencodelists.org/codelist/opensafely/systolic-blood-pressure-qof/3572b5fb/> | Minimum: 30; Maximum: 300 |
| eGFR | GDPPR | <https://www.opencodelists.org/codelist/nhsd-primary-care-domain-refsets/egfr_cod/20200812/> | Minimum: 0; Maximum: 200 |
| Glycated haemoglobin | GDPPR | <https://www.opencodelists.org/codelist/opensafely/glycated-haemoglobin-hba1c-tests/2ab11f20/> | Minimum: 3; Maximum: 250 |
| Total cholesterol | GDPPR | <https://www.opencodelists.org/codelist/nhsd-primary-care-domain-refsets/tcholhdl_cod/20200812/> | Minimum: 0.5; Maximum: 30 |
| HDL cholesterol | GDPPR | <https://www.opencodelists.org/codelist/nhsd-primary-care-domain-refsets/hdlcchol_cod/20200812/> | Minimum: 0.1; Maximum: 10 |
| Atrial fibrillation | GDPPR, HES-APC | [https://phenotypes.healthdatagateway.org/phenotypes/PH987/version/2165/detail/](https://conceptlibrary.saildatabank.com/phenotypes/PH987/version/2165/detail/) |  |
| Cancer | GDPPR, HES-APC | [https://phenotypes.healthdatagateway.org/phenotypes/PH960/version/2138/detail/](https://conceptlibrary.saildatabank.com/phenotypes/PH960/version/2138/detail/) |  |
| Dementia | GDPPR, HES-APC | [https://phenotypes.healthdatagateway.org/phenotypes/PH963/version/2141/detail/](https://conceptlibrary.saildatabank.com/phenotypes/PH963/version/2141/detail/) |  |
| Diabetes | GDPPR, HES-APC | [https://phenotypes.healthdatagateway.org/phenotypes/PH965/version/2143/detail/](https://conceptlibrary.saildatabank.com/phenotypes/PH965/version/2143/detail/) |  |
| Hypertension | GDPPR, HES-APC | [https://phenotypes.healthdatagateway.org/phenotypes/PH970/version/2148/detail/](https://conceptlibrary.saildatabank.com/phenotypes/PH970/version/2148/detail/) |  |
| Arrhythmia | HES-APC | [https://phenotypes.healthdatagateway.org/phenotypes/PH971/version/2149/detail/](https://conceptlibrary.saildatabank.com/phenotypes/PH971/version/2149/detail/) |  |
| Obesity | GDPPR, HES-APC | [https://phenotypes.healthdatagateway.org/phenotypes/PH976/version/2154/detail/](https://conceptlibrary.saildatabank.com/phenotypes/PH976/version/2154/detail/) |  |
| COPD | GDPPR, HES-APC | [https://phenotypes.healthdatagateway.org/phenotypes/PH991/version/2169/detail/](https://conceptlibrary.saildatabank.com/phenotypes/PH991/version/2169/detail/) |  |
| Angina | GDPPR, HES-APC | [https://phenotypes.healthdatagateway.org/phenotypes/PH956/version/2134/detail/](https://conceptlibrary.saildatabank.com/phenotypes/PH956/version/2134/detail/) |  |
| Hypercholesterolaemia | GDPPR | [https://phenotypes.healthdatagateway.org/phenotypes/PH1009/version/2187/detail/](https://conceptlibrary.saildatabank.com/phenotypes/PH1009/version/2187/detail/) |  |
| Depression | GDPPR, HES-APC | [https://phenotypes.healthdatagateway.org/phenotypes/PH1005/version/2183/detail/](https://conceptlibrary.saildatabank.com/phenotypes/PH1005/version/2183/detail/) |  |
| Deep vein thrombosis | GDPPR, HES-APC | [https://phenotypes.healthdatagateway.org/phenotypes/PH1007/version/2185/detail/](https://conceptlibrary.saildatabank.com/phenotypes/PH1007/version/2185/detail/) |  |
| Liver Disease | GDPPR, HES-APC | [https://phenotypes.healthdatagateway.org/phenotypes/PH1014/version/2192/detail/](https://conceptlibrary.saildatabank.com/phenotypes/PH1014/version/2192/detail/) |  |
| Chronic kidney disease | GDPPR, HES-APC | [https://phenotypes.healthdatagateway.org/phenotypes/PH990/version/2168/detail/](https://conceptlibrary.saildatabank.com/phenotypes/PH990/version/2168/detail/) |  |
| Charlson comorbidity index | HES-APC | <https://phenotypes.healthdatagateway.org/phenotypes/PH1658/version/3431/detail/> |  |
| COVID-19 | GDPPR, HES-APC, SGSS, Pillar 2, CHESS | <https://phenotypes.healthdatagateway.org/phenotypes/PH1/version/2/detail/> |  |
| Anticoagulants | Primary care dispensed medications | BNF codes beginning with: “0208” |  |
| Antiplatelets | Primary care dispensed medications | BNF codes beginning with: “0209” |  |
| Antihypertensives | Primary care dispensed medications | BNF codes beginning with: “0205” |  |
| Lipid lowering drugs | Primary care dispensed medications | BNF codes beginning with: “0212” |  |

Table S7: The RECORD statement – checklist of items, extended from the STROBE statement, that should be reported in observational studies using routinely collected health data.

|  | **Item No.** | **STROBE items** | **Location in manuscript where items are reported** | **RECORD items** | **Location in manuscript where items are reported (page numbers)** |
| --- | --- | --- | --- | --- | --- |
| **Title and abstract** | | | | | |
|  | 1 | (a) Indicate the study’s design with a commonly used term in the title or the abstract (b) Provide in the abstract an informative and balanced summary of what was done and what was found | Title indicates that health system data was used with specific datasets and that this was a retrospective cohort study mentioned in abstract. | RECORD 1.1: The type of data used should be specified in the title or abstract. When possible, the name of the databases used should be included.  RECORD 1.2: If applicable, the geographic region and timeframe within which the study took place should be reported in the title or abstract.  RECORD 1.3: If linkage between databases was conducted for the study, this should be clearly stated in the title or abstract. | Title & Abstract  (page 1)  Abstract  (page 1)  Abstract  (page 1) |
| **Introduction** | | | | | |
| Background rationale | 2 | Explain the scientific background and rationale for the investigation being reported | Introduction  (page 5) |  | Introduction  (page 5) |
| Objectives | 3 | State specific objectives, including any prespecified hypotheses | Introduction  (page 5) |  | Introduction  (page 5) |
| **Methods** | | | | | |
| Study Design | 4 | Present key elements of study design early in the paper | Methods  (page 6) |  | Methods  (page 6) |
| Setting | 5 | Describe the setting, locations, and relevant dates, including periods of recruitment, exposure, follow-up, and data collection | Methods  (pages 6-8) |  | Methods  (pages 6-8) |
| Participants | 6 | *(a) Cohort study* - Give the eligibility criteria, and the sources and methods of selection of participants. Describe methods of follow-up  *Case-control study* - Give the eligibility criteria, and the sources and methods of case ascertainment and control selection. Give the rationale for the choice of cases and controls  *Cross-sectional study* - Give the eligibility criteria, and the sources and methods of selection of participants  *(b) Cohort study* - For matched studies, give matching criteria and number of exposed and unexposed  *Case-control study* - For matched studies, give matching criteria and the number of controls per case | Methods  (pages 6-8) | RECORD 6.1: The methods of study population selection (such as codes or algorithms used to identify subjects) should be listed in detail. If this is not possible, an explanation should be provided.  RECORD 6.2: Any validation studies of the codes or algorithms used to select the population should be referenced. If validation was conducted for this study and not published elsewhere, detailed methods and results should be provided.  RECORD 6.3: If the study involved linkage of databases, consider use of a flow diagram or other graphical display to demonstrate the data linkage process, including the number of individuals with linked data at each stage. | Methods  (pages 6-8) |
| Variables | 7 | Clearly define all outcomes, exposures, predictors, potential confounders, and effect modifiers. Give diagnostic criteria, if applicable. | Phenotype definitions included in table "Table S6.a - Codelists and data sources used to define study variables and phenotypes" (page 36) | RECORD 7.1: A complete list of codes and algorithms used to classify exposures, outcomes, confounders, and effect modifiers should be provided. If these cannot be reported, an explanation should be provided. | Phenotype definitions included in table "Table S6.a - Codelists and data sources used to define study variables and phenotypes" (page 36) |
| Data sources/ measurement | 8 | For each variable of interest, give sources of data and details of methods of assessment (measurement).  Describe comparability of assessment methods if there is more than one group | Methods  (pages 6-8) |  | Methods  (pages 6-8) |
| Bias | 9 | Describe any efforts to address potential sources of bias | Methods (page 6) |  | Methods (page 6) |
| Study size | 10 | Explain how the study size was arrived at | Methods (page 6) |  |  |
| Quantitative variables | 11 | Explain how quantitative variables were handled in the analyses. If applicable, describe which groupings were chosen, and why | Statistical Analyses (page 7-8) |  |  |
| Statistical methods | 12 | (a) Describe all statistical methods, including those used to control for confounding  (b) Describe any methods used to examine subgroups and interactions  (c) Explain how missing data were addressed  (d) *Cohort study* - If applicable, explain how loss to follow-up was addressed  *Case-control study* - If applicable, explain how matching of cases and controls was addressed  *Cross-sectional study* - If applicable, describe analytical methods taking account of sampling strategy  (e) Describe any sensitivity analyses | Statistical Analyses (page 7-8) |  |  |
| Data access and cleaning methods |  | .. |  | RECORD 12.1: Authors should describe the extent to which the investigators had access to the database population used to create the study population.  RECORD 12.2: Authors should provide information on the data cleaning methods used in the study. | Methods  (pages 6-7)  Methods  (pages 6-7) |
| Linkage |  | .. |  | RECORD 12.3: State whether the study included person-level, institutional-level, or other data linkage across two or more databases. The methods of linkage and methods of linkage quality evaluation should be provided. | Abstract (page 1) & Methods (page 6) |
| **Results** | | | | | |
| Participants | 13 | (a) Report the numbers of individuals at each stage of the study (*e.g.*, numbers potentially eligible, examined for eligibility, confirmed eligible, included in the study, completing follow-up, and analysed)  (b) Give reasons for non-participation at each stage.  (c) Consider use of a flow diagram | Methods/Study population (page 6) | RECORD 13.1: Describe in detail the selection of the persons included in the study (*i.e.,* study population selection) including filtering based on data quality, data availability and linkage. The selection of included persons can be described in the text and/or by means of the study flow diagram. | Methods/Study population (page 6) |
| Descriptive data | 14 | (a) Give characteristics of study participants (*e.g.*, demographic, clinical, social) and information on exposures and potential confounders  (b) Indicate the number of participants with missing data for each variable of interest  (c) *Cohort study* - summarise follow-up time (*e.g.*, average and total amount) | Table 1 |  | Table 1 |
| Outcome data | 15 | *Cohort study* - Report numbers of outcome events or summary measures over time  *Case-control study* - Report numbers in each exposure category, or summary measures of exposure  *Cross-sectional study* - Report numbers of outcome events or summary measures |  |  | Table 1  Figure 4 |
| Main results | 16 | (a) Give unadjusted estimates and, if applicable, confounder-adjusted estimates and their precision (e.g., 95% confidence interval). Make clear which confounders were adjusted for and why they were included  (b) Report category boundaries when continuous variables were categorized  (c) If relevant, consider translating estimates of relative risk into absolute risk for a meaningful time period |  |  | Figure 1  Figure 2  Figure 3  Figure 5 |
| Other analyses | 17 | Report other analyses done—e.g., analyses of subgroups and interactions, and sensitivity analyses |  |  | Statistical Analyses (page 7-8) and STable1 |
| **Discussion** | | | | | |
| Key results | 18 | Summarise key results with reference to study objectives | Discussion  (pages 11-12) |  | Discussion  (pages 11-12) |
| Limitations | 19 | Discuss limitations of the study, taking into account sources of potential bias or imprecision. Discuss both direction and magnitude of any potential bias | Discussion  (page 12) | RECORD 19.1: Discuss the implications of using data that were not created or collected to answer the specific research question(s). Include discussion of misclassification bias, unmeasured confounding, missing data, and changing eligibility over time, as they pertain to the study being reported. | Discussion (pages 12) |
| Interpretation | 20 | Give a cautious overall interpretation of results considering objectives, limitations, multiplicity of analyses, results from similar studies, and other relevant evidence | Discussion  (pages 11-12) |  | Discussion (pages 11-12) |
| Generalisability | 21 | Discuss the generalisability (external validity) of the study results | Discussion (page 11) |  | Discussion (page 11) |
| **Other Information** | | | | | |
| Funding | 22 | Give the source of funding and the role of the funders for the present study and, if applicable, for the original study on which the present article is based | Funding information included in Acknowledgements  (page 13) |  | Acknowledgements (page 13) |
| Accessibility of protocol, raw data, and programming code |  | .. |  | RECORD 22.1: Authors should provide information on how to access any supplemental information such as the study protocol, raw data, or programming code. | Data Access/Methods |

*Reference: Benchimol EI, Smeeth L, Guttmann A, Harron K, Moher D, Petersen I, Sørensen HT, von Elm E, Langan SM, the RECORD Working Committee. The REporting of studies Conducted using Observational Routinely-collected health Data (RECORD) Statement. *PLoS Medicine* 2015; in press.

*Checklist is protected under Creative Commons Attribution ([CC BY](http://creativecommons.org/licenses/by/4.0/)) license.

### Figures

Figure S1 - Density distribution of the date difference in the earliest stroke record between data sources: GDPPR and HES-APC (n=188,970), GDPPR and SSNAP (n=155,220), and SSNAP and HES-APC (n=204,185).

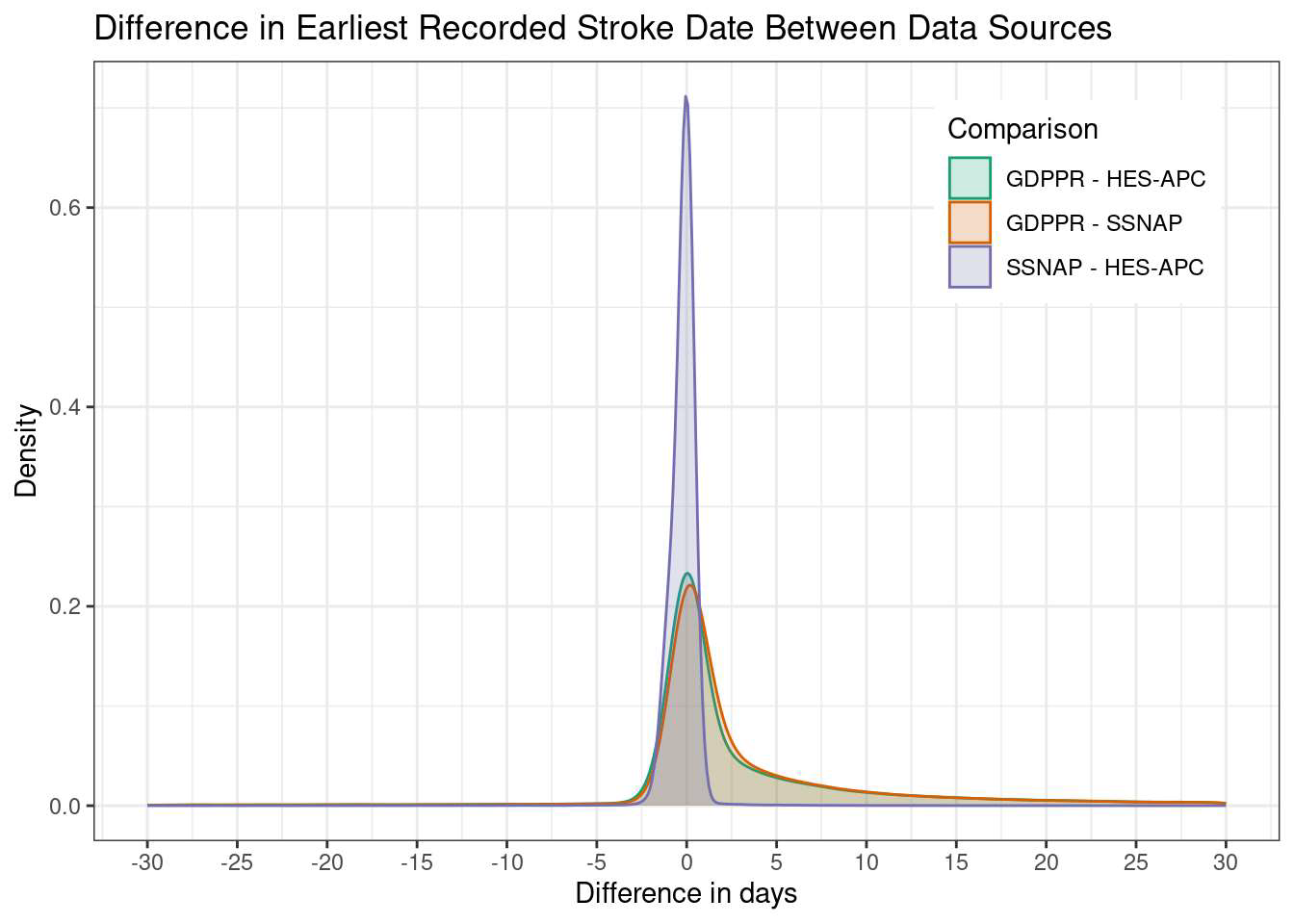

Figure S2 - Chord diagram illustrating the harmonisation of stroke type across 963,500 records from GDPPR, HES-APC, SSNAP, and ONS Deaths, corresponding to 425,675 individuals. Stroke type is assigned using a prioritisation approach, giving precedence to deterministic classifications from SSNAP, HES-APC, GDPPR, and ONS Deaths. Scale in thousands of records.

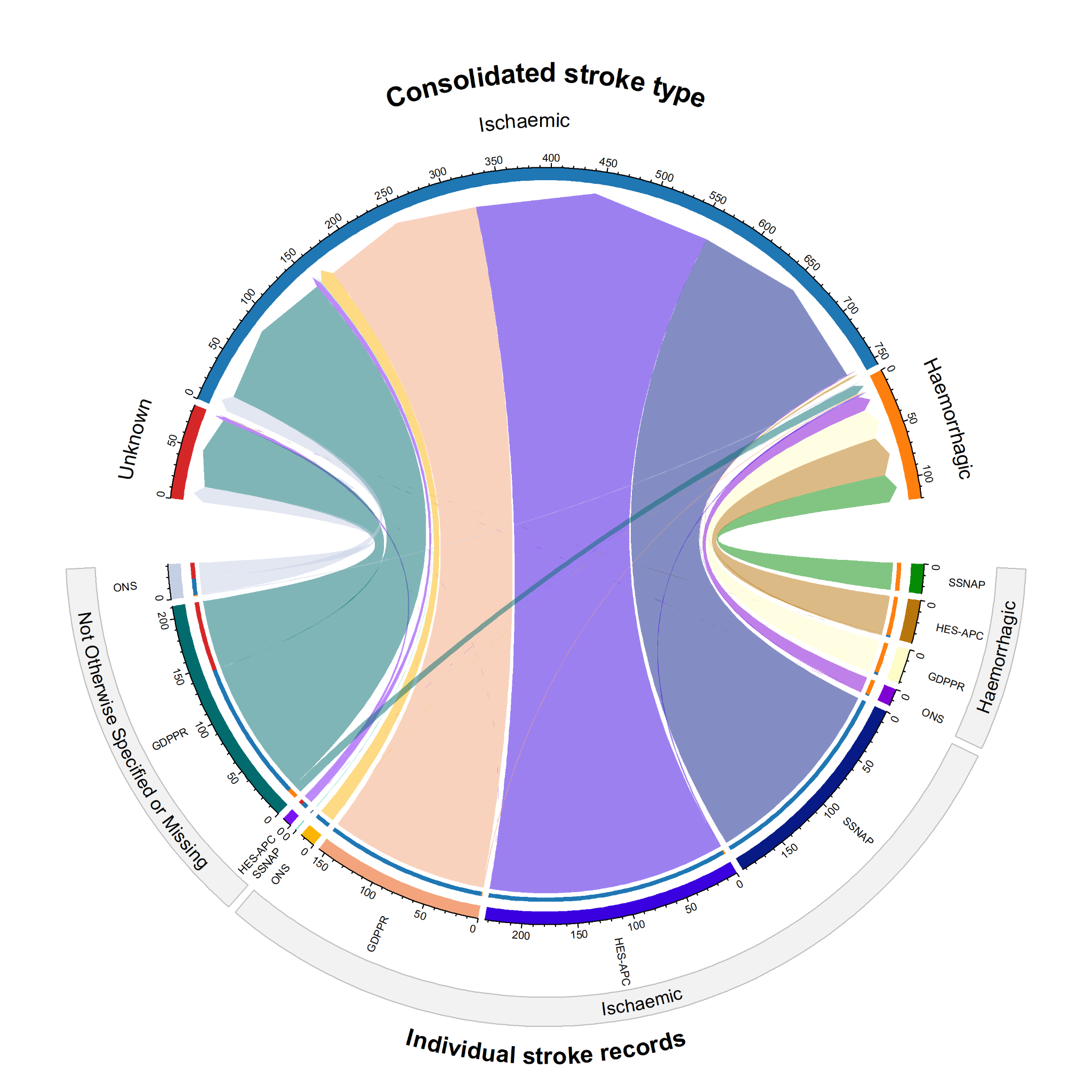

Figure S3a - One-year cumulative incidence of anticoagulants, antiplatelets, antihypertensives, and lipid-lowering drugs dispensed to individuals with non-fatal ischaemic strokes or unknown stroke type identified through GDPPR, HES-APC, or SSNAP (n=348,300).

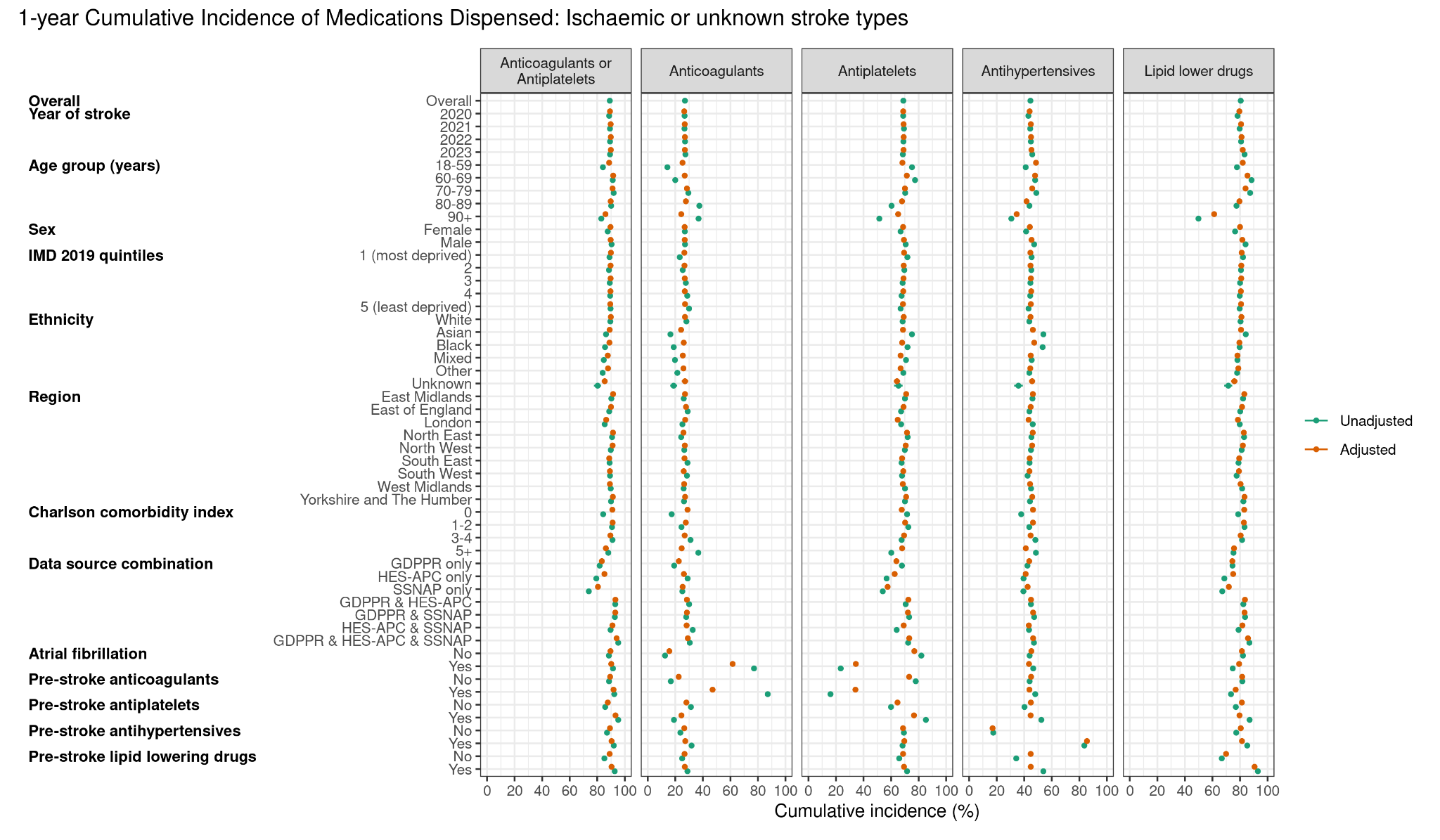

Figure S3b - One-year cumulative incidence of anticoagulants, antiplatelets, antihypertensives, and lipid-lowering drugs dispensed to individuals with non-fatal haemorrhagic strokes identified through GDPPR, HES-APC, or SSNAP (n = 34,435).

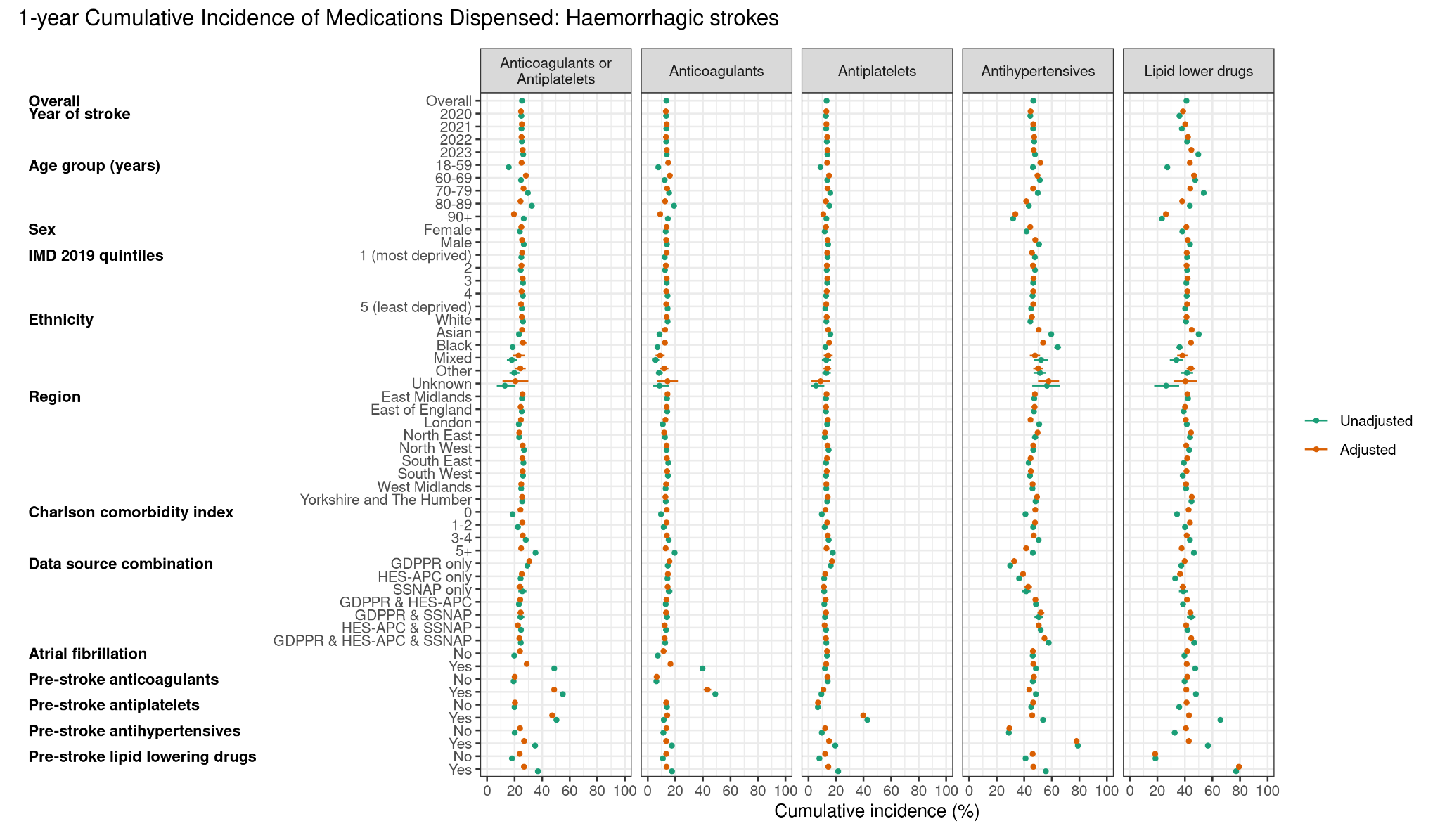
